# Markerless Motion Capture Reveals Movement Abnormalities in Isolated REM Sleep Behavior Disorder

**DOI:** 10.64898/2026.08.28.26361609

**Authors:** Philipp Wegner, Anja Ophey, Sinah Röttgen, Konstantin Kufer, Christopher E.J. Doppler, Aline Seger, Gereon R. Fink, Elke Kalbe, Komal Kotra, Marcus Grobe-Einsler, Katrin Feldmann, Michael Sommerauer, Jennifer Faber

## Abstract

Objective and scalable approaches for detecting subtle motor impairment in isolated REM sleep behavior disorder (iRBD), a prodromal stage of Parkinson’s disease, remain limited. We investigated whether markerless motion capture from single RGB-camera videos can identify gait abnormalities in people living with iRBD and provide interpretable digital biomarkers. We retrospectively analyzed 93 standardized walking videos from three clinical sites. Human pose estimation extracted 12 body markers and 14 kinematic time series. Thirty-five machine learning approaches classified healthy controls (HC) and people with iRBD. The Movement Disorder Society Unified Parkinson’s Disease Rating Scale Part 3 (MDS-UPDRS III) served as the clinical baseline. The best-performing model (tsfresh+XGBoost) achieved an AUROC of 0.739, significantly outperforming the MDS-UPDRS III sum score when trained on data from all three sites. Harmonized multi-site training improved performance. SHAP identified hip-related temporal features as key contributors, which differed between groups and showed stronger associations with regional dopaminergic deficits than clinical scores. Single-camera gait analysis may provide scalable digital biomarkers for low-cost screening and monitoring of prodromal PD.

**Plain Language Summary:** People with isolated REM sleep behavior disorder (iRBD) often show subtle changes in their walking that can be difficult to detect during routine clinical examinations. In this study, we used artificial intelligence to analyze videos of a simple walking task without requiring wearable sensors or body markers. The computer-based analysis distinguished people with iRBD from healthy individuals more accurately than standard clinician-rated motor scores. While further validation in larger studies is needed, these findings suggest that video-based movement analysis could become a useful objective tool to support the assessment of people at increased risk of Parkinson’s disease and related disorders.

## 1 Introduction

Early and reliable identification of persons at high risk for Parkinson’s disease (PD) is crucial for the development and implementation of neuroprotective interventions. PD is a neurodegenerative disease with movement abnormalities as a central symptom accompanied by non-motor impairment according to the Movement Disorders Society (MDS) [1]. A key prodromal marker for PD is isolated rapid eye movement (REM) sleep behavior disorder (iRBD) [2].

The characteristic loss of atonia during REM sleep leads to dream-enactment behaviors, such as violent movements, including kicking or hitting [3, 4]. While iRBD shows limited sensitivity at the population level, as only 33.3% of de novo individuals with PD report RBD symptom onset prior to motor manifestations [5], its value lies in defining a high-risk cohort. Approximately 80-90% of individuals affected by iRBD convert to either PD, Lewy body dementia (DLB), multiple-system atrophy (MSA), within 15 years after the initial iRBD diagnosis [6–8]. Misfolded α-synuclein is detectable in the cerebrospinal fluid and skin of individuals with iRBD, supporting its role as an early synucleinopathy [9–11]. Subtle motor abnormalities, either measured with clinical scales or quantitatively (e.g., sensor-based motor testing) have been described in prodromal PD and iRBD preceding clinical diagnosis for several years. [12–19] Particularly, motor changes were associated with higher rates of phenoconversion [15, 20].

Clinical rating scales such as the MDS Unified Parkinson’s Disease Rating Scale part III (MDS-UPDRS III, [21]) have been shown to exhibit increasing scores up to 5 years prior to a PD diagnosis and to achieve high specificity and sensitivity in the isolation of individuals with prodromal PD, however, their dependence on expert evaluation, along with floor effects and inter-rater variability at lower score ranges, may reduce sensitivity in early oligosymptomatic stages of the disease [12, 22–24]. In particular, such floor effects justify investigations for measures beyond established clinical scales that are potentially capable of modeling movement abnormalities in prodromal PD more fine-grained.

Quantitative assessment of motor impairment in PD has been extensively studied using wearable sensors, with multiple systematic reviews highlighting their widespread adoption and versatility for capturing motor symptoms such as gait, tremor, and bradykinesia [25–27]. In contrast, video-based approaches have only recently emerged and are underrepresented in the literature, and to our knowledge, single-camera markerless motion capture has not been applied to iRBD.

Besides alterations, e.g., in voice and finger tapping performance, gait abnormalities have been described in iRBD, suggesting movement analysis as a suitable tool to study motor impairments in individuals diagnosed with iRBD [18, 28–36].

Studies using wearable sensors with incorporated accelerometers or gyroscopes, multi-camera settings, or pressure plates have described several gait abnormalities in individuals with PD, including a decreased range of motion, increased variation in stride-length,-time, and-velocity, as well as increased asymmetry in arm swings [30–32, 37] In sensor-free, video-based approaches, full-body or zoomed-in videos of selected body parts are analyzed. These approaches are based on markerless motion capture, a computer vision paradigm that detects and tracks subjects (e.g., humans or animals) directly from video data without physical markers. Central to this paradigm are human pose estimation algorithms, typically deep neural networks trained to identify anatomical landmarks such as the knees, wrists, or nose in individual video frames and to track their trajectories over time [38–41]. In the context of movement quantification, the resulting time series (i.e., landmark trajectories over time) of extracted body markers serve as the basis for reconstructing movement trajectories and deriving quantitative kinematic parameters, including speed, amplitude, and variability of body positions, that allow a rater-independent characterization of individual motor patterns.

Video-based, markerless motion capture approaches are emerging as powerful tools for identifying and quantifying of movement and gait abnormalities in manifest PD and other neurodegenerative diseases. [33, 42–44]

In particular, such approaches require only minimal technical infrastructure that can even be reduced to a single monocular RGB camera [41, 42], and thus enable easily deployable, broadly applicable, and scalable data collection in clinical settings and even remotely in patients’ homes [45].

Early detection of motor impairments is critical, and previous studies have demonstrated gait alterations in individuals with iRBD compared to healthy controls (HC). Building on this evidence, the primary aim of the present study was to evaluate whether markerless motion capture applied to a simple walking task, recorded with a single front-view camera, provides sufficient information to distinguish iRBD from healthy controls. By leveraging an interpretable machine learning framework, we aimed not only to achieve accurate classification but also to elucidate the movement patterns that drive model outputs, thereby advancing our understanding of early motor changes in prodromal neurodegeneration. Furthermore, we sought to determine how such video-derived markers relate to established clinical rating scales, such as the MDS-UPDRS III, and to dopaminergic imaging (DaT-SPECT), an established marker of neurodegeneration associated with higher rates of phenoconversion to PD. [46–49].

Based on prior evidence of subtle motor changes in iRBD, we hypothesized that even a minimalistic video setup—using a single front-view camera and a simple walking task would be sufficient to capture clinically relevant motor alterations distinguishing iRBD from HC.

Video data were retrospectively collected at three sites, with an unbalanced distribution of iRBD and HC across sites. State-of-the-art markerless motion capture models were used to extract 12 body landmark positions from each video frame, yielding multivariate time series. We evaluated different time-series machine learning (ML) approaches for distinguishing iRBD from HC and addressed the site imbalance using a fixed test set collected at a single site, so that group membership was not confounded with recording site. Both supervised and unsupervised learning paradigms were considered.

To improve the interpretability of the classification results, we subsequently identified time-series features that contributed most strongly to group separation, providing insights into the underlying motor behavior. Finally, in a subgroup of individuals with iRBD, we investigated the clinical relevance of these features by correlating them, alongside MDS-UPDRS III scores, with regional z-scores for dopaminergic uptake measured by DaT-SPECT.

## 2 Methods

### 2.1 Data

#### 2.1.1 Participants

Data of this retrospective study included data that were collected across three sites in Germany, referred to here as site A, B, and C, from participants of observational studies (i) diagnosed with isolated REM sleep behavior disorder (iRBD) by overnight videopolysomnography (vPSG) [50] from sites A and B, and (ii) healthy controls (HC) from sites A and C. Sites A and B contributed data as part of a cohort study on movement disturbances in iRBD [50, 51] (Trialnumber: DRKS00024898), while site C provided data of HC from a cohort study on neurodegenerative ataxias that employed a matching experimental gait protocol (Trialnumber: DRKS00008304). Demographic information on age and sex was available for all participants. MDS-UPDRS III motor score was available for all individuals with iRBD from site A and B, and HC from site A. In addition, diagnostic DaT-Scan results were available in a subset of individuals with iRBD from site A. All participants gave informed consent, and the observational studies were approved by the respective ethics committees (https://medfak.uni-koeln.de/forschung/translation-i-klinische-forschung/ethikkommission, https://ethik.meb.uni-bonn.de/).

#### 2.1.2 Gait task

All participants performed a standard walking task that was videotaped using a single monocular RGB camera as illustrated in Figure 1 (I). The entire walk, including both outbound and return segments, was used in the analysis. The walking task with simultaneous video recording was conducted as follows: Each participant was instructed to start at a fixed starting position, facing away from the camera (Figure 1, (I) Point A). From there, the participant walked in a straight line at a natural, everyday walking pace toward a predefined point at least 6m away aligned with the camera’s viewing direction (Figure 1, Point B). Upon reaching Point B, the participant executed a 180*^◦^* turn and returned along the same straight path to the starting position, maintaining a self-paced, comfortable walking speed.

**Figure 1.**
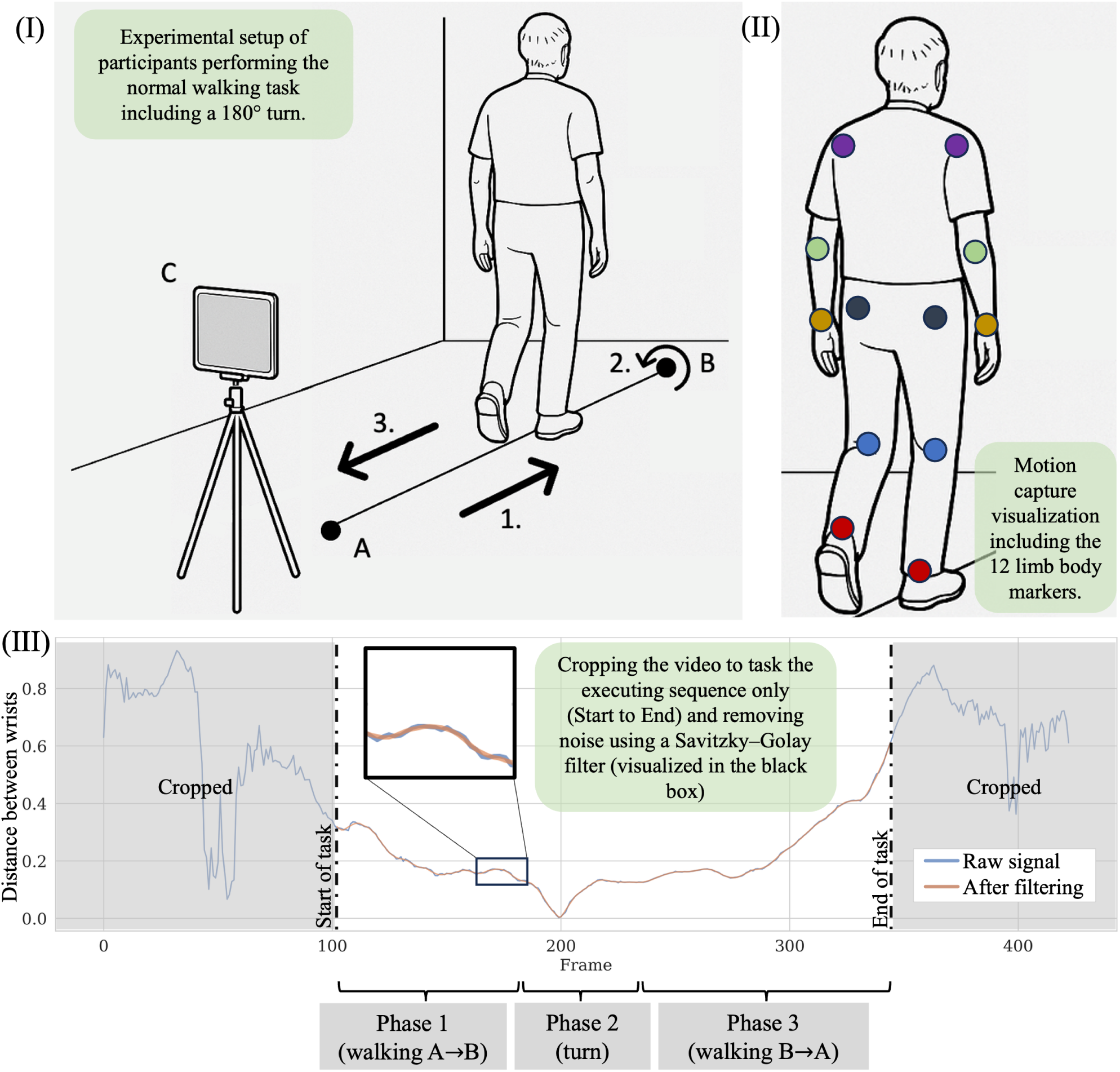
Illustration of the clinical setup and exemplary time-series data extracted from markerless motion capture. (I) Within the video-captured gait task, each participant first walks in a straight line from the starting point (A) to the turning point (B) at a self-selected, comfortable walking pace; second, performs a 180*^◦^* turn, and third, walks back to the starting point (A). The RGB camera (C) is aligned with the walking path 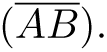 (II) Using a human pose estimation model, a movement skeleton comprising 12 body markers representing the joints of the upper and lower limbs is extracted from each video frame of the videotaped gait task. (III) The pose estimation outputs are preprocessed before being fed into the downstream analyses. Each video was visually inspected for quality control and manually labeled with the start and end frames of actual task execution with full-body detection, excluding, if present, the initial and final video segments during which participants position themselves at the starting point or leave the scene after task execution. A Savitzky–Golay filter was applied to reduce noise arising from the discrete temporal sampling (frames per second), which introduces slight jitter in body marker positions from frame to frame. A representative time series of the pose estimation output is shown before (blue) and after filtering (orange), with a zoomed-in inset, illustrating the filtering step. The orange plotted time series represents the data as in the downstream analyses.

#### 2.1.3 Video-data acquisition and quality control

All videos were captured at a 1920×1080 pixel spatial resolution, with a temporal resolution of 30 or more frames per second (fps). Each video was visually inspected for quality control to check usability. Criteria for the exclusion of videos included falsely executed walking tasks and environmental factors that could potentially influence the participant or the videotaping, e.g., disturbance of the overall scene by other persons.

### 2.2 Computer vision-based spatiotemporal motion quantification

Each suitable video underwent a structured processing pipeline as follows.

#### 2.2.1 Motion capture

First, motion capture was conducted using the YOLO11 pose estimation model [52] to extract the standard set of 17 body markers defined in the COCO dataset [53]. All 12 body markers representing upper and lower limb joints, namely left and right shoulder, elbow, wrist, hip, knee, and ankle, were used for the downstream analysis in this work. The remaining 5 head markers (left and right eye and ear, and the nose) were not included. The per-frame motion capture output is visualized in Figure 1 (II), in which a person is overlaid with the 12 included body markers. These markers were assessed irrespective of whether the person is facing the camera or not.

#### 2.2.2 Pose estimation output preprocessing

The 12 pose estimation output signals were further preprocessed. First, each video was manually cropped to the actual execution of the walking task with full-body coverage. Where necessary, initial and final frames (e.g., capturing movement toward the starting position or the subject leaving the scene) were excluded. The start frame was set to the first frame of walking task execution in which all 12 body markers were successfully detected. The end frame was defined as the last frame in which all 12 body markers were successfully detected. This manual step was performed blinded to the groups, iRBD vs. HC. Second, the pose estimation output from the remaining task-related part of the videos, sampled at > 30 fps, was downsampled to ensure a homogeneous sampling rate of 30 fps across all subjects. Third, missing pose estimation outputs were imputed by linear interpolation. During the walking task, particularly while turning, individual body landmarks were occasionally occluded and therefore not detected by the markerless motion capture algorithm in certain video frames. Missing landmark coordinates were subsequently reconstructed using linear interpolation between the nearest available observations. Fourth, a Savitzky–Golay filter (window length = 11, polynomial order = 3) was applied. As pose estimation operates frame-wise, discrete temporal sampling results in a discontinuity in the signal from frame to frame. Filtering was applied to mitigate this data’s inherent noise while preserving signal dynamics (Figure 1(III)).

#### 2.2.3 Time series extraction

Next, from these preprocessed 12 pose-estimation signals, multiple time series describing kinematic relationships among body markers were extracted. In total, 14 distinct time series per recording were included in the downstream processing and analyses, comprising (i) pairwise distances between body markers (e.g. the distance between both ankles), (ii) joint angles defined by triplets of body markers (e.g. left shoulder angle with apex at the left shoulder and basis formed by left hip and wrist), and (iii) areas defined by quadruplets of body markers (e.g., the rectangle formed by both hips and both ankles). The selection of the 14 time series was based on prior work [54]. A detailed list of all included time series is provided in the Supplementary Material (Suppl. Table S1).

### 2.3 ML Model Comparison: Experimental Design and Evaluation Strategy

We compiled different supervised and unsupervised machine learning (ML) models and compared them with respect to their ability to distinguish movement patterns in iRBD from those of healthy controls (HC), framed as a classification problem. Compilation of training and test sets took into account the imbalanced distribution of classes across sites. Hence, the test set throughout all experiments remained the data from site A. The experimental design and evaluation strategy for each respective ML model architecture and training set compilation are described in the following section. Before undertaking the actual predictive modeling, a logistic regression was conducted to investigate the potential confounders age, sex, and video length that might confound with class, and thus could lead to shortcut learning (for more details see Supplementary Data, Section S3). Throughout this section, “experiment” or “experimental design” refers to a specific combination of a training set configuration and ML model architecture applied to solve the iRBD vs. HC classification task.

#### 2.3.1 Compilation of Training Sets and Subsequent Experimental Design

Training was performed using single-site data (site A comprising both classes, site B comprising iRBD only, site C comprising HC only) and combinations of two or all three sites. This resulted in seven training set compositions, ranging from clean single-site data comprising both classes (iRBD and HC; Site A) to enriched combinations (sites A+B, A+C, A+B+C) and fully site-confounded settings (sites B, C, B+C). The imbalanced multi-site dataset introduced potential site-dependent bias while also enabling assessment of whether unbalanced, enriched datasets can nevertheless enhance model performance. To mitigate the risk of site-related shortcut learning and biased performance evaluation, the experimental design included the following four strategies: (i) All ML models were tested on the single-site dataset, which comprised both classes (Site A) to prevent site–class confounding and ensure site-unbiased performance estimates. (ii) For all experiments where Site A data was part of the training set (Site A only, Site A + B/C/B+C), the model evaluation was performed using a leave-one-out cross-validation (LOOCV) on Site A. In each fold, a single sample from Site A was held out for testing, while the remaining Site A samples, and in multi-site training data compositions, all data from the additional training sites (Sites B and/or C), were used for training. Accordingly, this cross-validation step was not applied to experiments with training data that excluded Site A, namely B, C, or B+C. Here, evaluation remained on the hold-out single-site A test set. (iii) For enriched training data sets (Site A + B/C/B+C), a cross-site harmonization was applied before training to align input data across sites in a class-aware way (B and C aligned to the respective class training fold of A) and reduce potential site effects. Detailed information on this class-aware cross-site harmonization of the training data is provided in Paragraph 2.3.2 - *Class-aware cross-site harmonization* after introducing the model architectures, since cross-site harmonization is tailored to the respective model inputs. (iv) To avoid a bias resulting from unevenly distributed numbers between classes, such class imbalances were resolved by augmentation of the minority class in the respective training data set. Detailed information on this class-imbalance correction of the training data is provided in Paragraph 2.3.2 - *Class-imbalance correction* after introducing the model architectures, since the class-imbalance correction is tailored to the respective model input data.

(v) To obtain robust and unbiased performance estimates, all experiments were repeated 30 times with different random initializations, i.e., different random seeds. Accordingly, the model performance metrics (see Subsection 2.3.5) for the classification experiments reported in this work are provided as the mean, standard deviation, and 95% confidence interval across these repetitions.

Depending on the composition of the respective training sets, either supervised ML models for the five training sets containing both classes (A, A + B/C/B+C, B+C), or unsupervised approaches for the two single-class training sets (C, B), were applied. Depending on the ML model architecture, either raw time series or features extracted from them served as input. All ML model architectures are described in detail in the following Subsections (2.3.2 and 2.3.3). All experiments are summarized in Table 1.

**Table 1:** Overview of experimental configurations used in this study and the respective learning paradigm. The left column lists all considered training site configurations, adding the respective learning paradigms that were applied in that given scenario. Different machine learning (ML) models were assessed for their ability to distinguish movement patterns in patients with isolated REM sleep behavior disorder (iRBD) from those of healthy controls (HC) using markerless, video-based motion capture, and framing the problem as a classification task. For one-class training set configurations, five different unsupervised ML methods were applied and evaluated on their performance in abnormality detection. For training set configurations comprising both classes, five different supervised ML models were applied and evaluated on their ability to distinguish iRBD from HC. To mitigate the risk of site-related shortcut learning and biased performance evaluation, the experimental design included several strategies, e.g., class-aware cross-site harmonization and class-imbalance correction, and all supervised and unsupervised models were evaluated exclusively on Site A, representing the only site with representation of both classes, to ensure a consistent test set across all experiments, thereby enabling comparability.

| Train Sites | Method Type | Approach |
| --- | --- | --- |
| B | Unsupervised | Abnormality detection |
| C | Unsupervised | Abnormality detection |
| B + C | Supervised | Classifier on perfectly site-confounded data |
| A | Supervised | Classifier baseline |
| A + B | Supervised | Classifier, training set enriched with all available iRBD data |
| A + C | Supervised | Classifier, training set enriched with all available HC data |
| A + B + C | Supervised | Classifier, training set enriched with more iRBD and HC data using all available data |

#### 2.3.2 Supervised Architectures

For the supervised learning setting, five different modeling strategies were evaluated. Two model architectures (S1, S2) utilized the 14 time series (Suppl. Table S1) stacked into one vector; three model architectures used feature sets extracted from the 14 time series (S3-5), thus resulting in a total of five distinct model types. To account for site effects and class imbalances, class-aware cross-site harmonization was applied to all enriched training data sets (A + B/C/B+C) and class-imbalance correction was achieved by augmenting the minority class in each training data set. Both mitigation strategies were tailored to the respective model input data of either raw time series (S1, S2) or feature sets extracted thereof (S3-5) and are described in more detail in paragraph 2.3.2 - *Class-aware cross-site harmonization* and paragraph 2.3.2 - *Class-imbalance correction*. Finally, each of these five model types results in a classifier. (S1) The first approach operated directly on the multivariate time series constructed by stacking the 14 individual time series as model input data without any feature extraction. After harmonization and augmentation, pairwise distances between participants were computed using dynamic time warping (DTW) [55], and a k-nearest neighbors (kNN) [56] classifier was applied to assign class labels based on these distances. (S2) The second approach employed the (Mini-)ROCKET framework [57], which transforms the input by applying 10,000 fixed random convolutional kernels to the stacked multivariate time series, as in (S1). The resulting feature representations were used to train a logistic regression classifier. Unlike the original (Mini-)ROCKET, which relies on ridge regression, logistic regression was chosen here because it provides class probability estimates, necessary to compute the AUROC and precision at 80% recall metrics. (S3-5) For each recording, the 14 time series were independently processed using three different feature extraction tools, namely (i) tsfresh [58], (ii) tsfel [59], and (iii) catch22 [60], to compute descriptive time-series features, including statistical moments, frequency-domain descriptors, and autocorrelation measures. The resulting feature sets from all 14 time series were concatenated to form a single fixed-length feature vector for each of the three feature-extraction tools. This can result in a high-dimensional feature vector, where the number of features may exceed the number of training samples. Accordingly, features were selected using a Random Forest–based approach [61], ranking them by their importance for the classification task (iRBD vs. HC). Only the n subsample% top-ranked features were retained, augmented, and passed to the downstream XGBoost model [62], thereby reducing the feature dimensionality to below the number of samples. The number of features retained was a tunable hyperparameter. This feature selection step was performed solely on the training set to avoid data leakage. Each combination of tsfresh+XGBoost (S3), tsfel+XGBoost (S4), and catch22+XGBoost (S5) was evaluated as a separate model type.

### Class-aware cross-site harmonization

Potential site-related differences, arising, e.g., from slightly varying camera setups or environments, might introduce distributional shifts that in turn can confound model training and evaluation. For enriched training data sets (Site A + B/C/B+C), a cross-site harmonization was applied prior to training to align input data across sites in a class-aware way (B and C aligned to the respective class training fold of A) and reduce potential site effects. To account for this risk, class-aware cross-site harmonization strategies were implemented, tailored to the respective type of input data:

*(i) Class-aware cross-site harmonization of input data type: Time series* For models operating directly on raw time series (S1, DTW–kNN, and S2-(Mini-)ROCKET), harmonization was performed using dynamic time warping (DTW)-based temporal alignment. In each experiment, subjects from the single-class sites B and C were matched to the most similar subject of the same class from the Site A training fold based on the minimum DTW distance. Importantly, matching was performed independently within each training fold to prevent information leakage. The optimal DTW alignment path obtained between the matched subjects was then used to warp the 14-dimensional time series of the target subject onto the temporal structure of the Site A reference subject.

*(ii) Class-aware cross-site harmonization of input data type: Feature sets extracted from time series* Harmonization was applied to each feature set provided by the three utilized feature extraction toolkits, namely tsfresh, tsfel, and catch22, to reduce domain shift due to the site while preserving the decision boundaries expected by the subsequently applied tree-based classifiers, in our case XGBoost. In each experiment, harmonization was performed on a per-feature basis, tailored to the corresponding training dataset, explicitly excluding the test set to avoid data leakage during harmonization. In detail, both single-class sites (Site B and C) were aligned to the corresponding class-specific Site A training fold. As an example, for the training set comprising sites A+B+C, since Site C contains only healthy controls (HCs), its features were shifted and rescaled to match the distribution of HCs in the training fold of the Site A data. Analogously, features from Site B, which comprises only individuals with iRBD, were aligned to the distribution of iRBDs included in the training fold of Site A. For more details, see Suppl. Section S2.

Notably, the training data from Site A remained unaltered, with only the single-class site data (Site B and Site C) were projected into the feature space defined by the clean Site A comprising both classes. This design was chosen to ensure that model performance reflects generalization under controlled domain adaptation rather than being confounded by site-specific artifacts. Moreover, class-aware cross-site harmonization of model inputs was performed separately for each experiment and, when applicable, within each cross-validation training fold. Harmonization was based exclusively on the corresponding Site A training fold, while the test set was explicitly excluded to prevent data leakage.

### Class-imbalance correction

Class imbalance between iRBD and HC in training sets was addressed by oversampling the minority class in each supervised experiment. This procedure was tailored to the different input datatypes used. For the first and second (S1, S2) modeling strategies, which used raw time series as input data, augmentation had to be aligned to the original temporal constraints only. Accordingly, the minority class was augmented by adding Gaussian noise to its samples until class balance was achieved. In the last modeling strategies (S3-5) utilizing time series features, feature spaces are non-uniform and specific for the respective features. Accordingly, the class-imbalance correction was performed using a separate algorithm, namely BorderlineSMOTE [63, 64], for each feature to ensure it remained within the feature space boundaries.

In summary, for the supervised experiments, each training fold underwent the following procedure. Initially, for enriched training samples, the entire training fold is harmonized onto the site A data, i.e., the site that holds both classes. Secondly, for modeling strategy S(3-5), namely *{*tsfresh/tsfel/catch22*}* + XGBoost, feature reduction was applied, with features sub-selected using Random Forest-based feature importance. Third, the minority class is augmented with approaches tailored to the specific input data type (either raw time series or time series features).

#### 2.3.3 Unsupervised Architectures

For all unsupervised approaches, models were trained on a single-class reference dataset (Site B or Site C) and evaluated on Site A. When trained on Site B (iRBD only), subjects from Site A were assessed relative to the iRBD distribution, whereas when trained on Site C (HC only), subjects were assessed relative to the HC distribution. This yields a subject-specific abnormality score for each test sample, depending on the corresponding reference class.

Five different modeling strategies were evaluated, with two approaches operating directly on the raw 14 time series (U1, U2), and three approaches based on engineered feature representations (U3-U5), resulting in five distinct model configurations.

(U1) Each participant is defined by its set of 14 time series (Suppl. Table S1), represented as a 14-dimensional multivariate time series. Pairwise distances between participants were computed using dynamic time warping (DTW) [55] applied to these representations. To quantify class-specific proximity, we used a k-nearest neighbour (kNN) approach in a cross-dataset setting. Specifically, all subjects from Site A (containing both classes) were evaluated against the two reference datasets: Site B (iRBD only) and Site C (HC only). For each subject in Site A, k nearest neighbours were identified separately in Site B and in Site C based on DTW distances. For each reference class, we computed the average DTW distance between the subject and its k nearest neighbours, yielding dissimilarity scores for each subject of A (A*→*B and A*→*C). The resulting scores were z-score normalized across subjects in Site A and mapped to the unit interval [0, 1] using a sigmoid transformation. A threshold of 0.5 was then applied to assign class labels, with subjects classified as iRBD or HC based on which reference class yielded the lower normalized distance. This procedure was performed separately for sites B and C.

(U2) The second approach applied 10,000 random convolutional kernels to the augmented 14 raw time series per participant, following the (Mini-)ROCKET framework [65]. The resulting feature representations were used to train a one-class support vector machine (one-class SVM) [66], which learns a decision boundary in the feature space that encloses most of the training samples (inliers). Samples lying outside this boundary are considered outliers.

(U3-U5) Three distinct feature sets derived from tsfresh, tsfel, and catch22, respectively, identical to those used in the supervised experiments (S3-5), were constructed. Each feature set was independently used to train an isolation forest [67] for outlier detection.

#### 2.3.4 Hyperparameters

Feature-based, supervised experiments (*{*tsfresh, tsfel, catch22*}* + XGBoost) included a dimensionality reduction step to mitigate overfitting, with dimensionality n a tunable hyperparameter. For XGBoost models, additional tunable hyperparameters included the learning rate, maximum tree depth, number of boosting rounds, and subsample rate. These were optimized with optuna (v4.4.0) in 30 trials per fold in the inner loop of the LOOCV. Because tuning occurred within the cross-validation loop, no single universal set of hyperparameters can be reported. For reproducibility, Supplementary Table S2 lists the average hyperparameters for an example experiment (tsfresh + XGBoost). The dynamic time warping+kNN was implemented with k = 2 and distance-based weighting. (Mini-)ROCKET combined with logistic regression was tuned for regularization strength using built-in 5-fold cross-validation in scikit-learn. Unsupervised detection experiments used one-class SVM (ν = 0.1), isolation forest, and dynamic time warping distances, with the latter two approaches applied using default parameters.

#### 2.3.5 Evaluation of model performance

As described above, model performance, defined as the ability to distinguish movement patterns observed in iRBD versus HC, was consistently evaluated on Site A data, which comprises the only data from both classes. Each experiment was repeated with 30 different random initializations to assess stability and generalizability. Performance metrics are therefore reported as the mean, standard deviation, and 95% confidence interval across these repetitions for each model. The supervised experiments in this work were evaluated using balanced accuracy and the macro-averaged F_1_-score, as well as area under the receiver operating characteristic curve (AUROC) and the maximal precision given a minimum recall of 80% (Precision@80%Recall). These metrics were selected to provide complementary perspectives on classification performance. AUROC quantifies a model’s overall ability to discriminate between classes across all possible decision thresholds. Precision@80%Recall evaluates performance under a clinically relevant operating condition by measuring the highest achievable precision while maintaining a recall of at least 80%. This metric was chosen to reflect the priorities of a potential screening application, where high recall is required to minimize missed cases, while high precision helps reduce false-positive findings. The calculation of Precision@80%Recall is illustrated in Supplementary Figure S2.

The unsupervised experiments were evaluated separately because the underlying model architectures do not inherently produce class probabilities, thereby precluding the direct computation of probability-based metrics such as AUROC. Performance was assessed using balanced accuracy and the macro-averaged F_1_-score.

For the overall best-performing model, a comprehensive in-depth evaluation was conducted by additionally reporting the weighted F_1_-score, the area under the precision-recall curve (AUPRC), and balanced accuracy.

### 2.4 Additional Analyses

#### 2.4.1 Comparison with clinical rating scale

To establish a human baseline for the HC vs. iRBD classification task, we evaluated human-rated motor scores according to the MDS-UPDRS rating scale. The MDS-UPDRS was conducted and videotaped by outcome assessors and rated by one of two movement disorder specialists (CEJD, AS) blinded for the diagnostic group (HC vs. iRBD). The summed MDS-UPDRS Part III motor score was used in a logistic regression model, which was evaluated using leave-one-out cross-validation on Site A, comprising both HC and individuals with iRBD.

Performance was assessed using the AUROC, Precision@80%Recall, and the macro-averaged F_1_-score. The best-performing clinical model served as the human baseline, providing a clinically grounded and methodologically comparable benchmark for the machine learning models. One-sided one-sample Wilcoxon signed-rank tests were used to compare the performance of the best-performing machine learning models against the human baseline. Statistical significance was defined as p < 0.05.

#### 2.4.2 Model-guided feature analysis

To gain insight into the pattern of the most relevant computer-vision-derived parameters and whether they might inform further downstream analyses, an explainable AI approach was implemented. This enabled ranking the most influential features and, accordingly, time series and body parts for differentiating iRBD from healthy controls, as well as further exploration of temporal dependencies in greater detail.

### SHAP analyses to identify time series and body parts of highest impact

SHapley Additive exPlanations (SHAP) [68] values were computed for each held-out test sample during the leave-one-out cross-validation (LOOCV) in the supervised experiment models S3-5 as outlined in Subsection 2.3.1), and subsequently averaged across all test samples, i.e., all samples from Site A. For the best-performing ML model type trained on the optimal training dataset, SHAP values for features from the 14 time series were calculated and ranked by their average across all LOOCV folds. For visualization, the three highest-ranked features were plotted in pairwise combination across three two-dimensional scatter plots. A support vector machine (SVM) with a sigmoid kernel was used to generate class probability overlays for each feature pair. This was done to illustrate the separability of the classes within the selected feature space and to visualize the corresponding decision regions learned by the model. Moreover, SHAP values for time series features were aggregated by body part and then averaged. Here, each time series was assigned to all body parts included in that time series. For instance, the bilateral distance time series *dist left wrist–left hip*) was attributed to both involved body parts, namely wrists and hips. Since body parts differ in the number of time series in which they are included, and therefore contribute unequally many features to the model, the mean rather than the sum of SHAP values was computed per body part. This ensures that body parts represented by more time series (e.g., hips, which appear in multiple distance and angle features) are not artificially inflated in their apparent importance. Body parts were then ranked by their mean SHAP value across all LOOCV folds.

### Downstream evaluation of temporal autocorrelation

SHAP analysis revealed that features encoding temporal dependencies contributed most strongly to the best-performing model (see Section 3.4). This motivated a more detailed investigation of partial autocorrelation functions (PACF) to further investigate the temporal characteristics of the most influential time series of the best-performing model. PACFs quantify the direct temporal dependency between time series values of the current frame (frame*_t_*) and the respective lag k earlier frame (frame*_t−k_*), while controlling for intermediate lags (1,…,k *−* 1) [69]. PACF values were estimated for lags 1–30 to compare the strength of direct temporal dependencies across lag distances between groups. More specifically, |PACF*_T_ |*: 1,…,30 *→* R with k *’→ |*PACF (T, k)|, where T is a time series. For more details on this term: PACF (T, k), see Supplementary Material S6. Downstream analysis in this work concerns only one specific time series; hence, the notations PACF*_T_* and PACF are equivalent, and the latter one is used here for simplicity. For each lag, |PACF| values were compared between individuals with iRBD and healthy controls using a two-sided Mann-Whitney U-test, resulting in 30 comparisons (one for each lag). Multiple comparisons were corrected using Benjamini-Hochberg. The error bars indicate the standard deviation for each lag per group. iRBD = group of individuals with iRBD; HC = healthy controls.

#### 2.4.3 Association with Dopaminergic Imaging

The subgroup of individuals with iRBD coming from site A, with available diagnostic DaT-Scan imaging results, enabled us to investigate the extent to which computer-vision-derived digital parameters and clinical measures relate to the underlying neurodegenerative pathology. Regional binding ratios from DaT-Scan imaging, reflecting dopaminergic deficits, were related to (i) the set of top 30 computer vision–derived features of the best-performing ML model, ranked as most influential by the explainable AI-informed SHAP analyses (TOP30, see Section 2.4.2), (ii) PACF of lag 1-30, representing the further exploration of the highest impact feature of the best-performing ML model (PACF30, see Section 2.4.2), and (iii) the best-performing clinical measure (see Section 2.4.1). Regional binding ratios (DaT-Scan z-scores) were assessed in the following eight brain regions: caudate, anterior/posterior/total putamen, each in the left and right hemisphere separately.

### UMAP-based derivation of scalar digital gait markers from computer-vision features

Both ((i) + (ii)) computervision-derived digital parameters are high-dimensional. Thus, these high-dimensional representations must be condensed into a single scalar value to enable analogous correlation and interpretation of those of the clinical scale. Uniform Manifold Approximation and Projection (UMAP) was applied to achieve the dimensionality reduction while preserving the underlying structure of the high-dimensional data in the low-dimensional, scalar representation [70]. First, the hyperparameter (n neighbors) needs to be defined. To prevent data leakage, the hyperparameters (n neighbors) were tuned on the independent “tuning set” of subjects from whom DaT-Scan imaging data were not available. Second, the optimal parameter was selected based on a composite score combining manifold trustworthiness (weight 0.7) and stability (weight 0.3) [70, 71]. The fitted UMAP models were subsequently applied to the DaT-Scan cohort to obtain, for each subject, a single scalar representation, which we refer to as TOP30 and PACF30, respectively. Note that mapping a vector into a lower-dimensional space might be referred to as an “embedding” in ML literature and accordingly also in this work.

### Association of regional dopaminergic imaging with clinical and computer-vision-derived measures

We assessed the association between regional dopaminergic binding and clinical rating scores, digital motor measures, and their combination using Ordinary Least Squares (OLS) linear regression [72]. All models were adjusted for age, sex, the time interval (in days) between the video assessment and clinical rating, and the time interval between the clinical assessment and DaT-Scan imaging.

For each of the eight brain regions assessed by DaT-Scan imaging, five regression models with different predictor sets were fitted to the respective regional binding ratio: (i) a clinical model including the best-performing clinical scale; (ii) a digital motor model including the TOP30 embedding; or (iii) a digital motor model including the PACF30 embedding; and two combined models including both the best-performing clinical measure and either (iv) the PACF30 embedding (PACF30 Combined) or (v) the TOP30 embedding (TOP30 Combined). The aforementioned covariates were included in all regression models.

To estimate model fit and its uncertainty, we employed a nonparametric bootstrap procedure with n = 1, 000 iterations. In each iteration, the dataset was resampled with replacement and all regression models were refitted. Model fit was assessed using the coefficient of determination (R^2^), the Akaike Information Criterion (AIC), and the corrected Akaike Information Criterion (AIC_c_) [73, 74]. For each model, 95% bootstrap confidence intervals (CIs) were calculated from the 2.5th and 97.5th percentiles of the corresponding bootstrap distributions. To compare model fit, the difference in R^2^ between models was calculated within each bootstrap iteration, thereby preserving the paired structure of the bootstrap samples. The resulting bootstrap distributions of ΔR^2^ were summarized by their point estimates and 95% CIs.

### 2.5 Code Availability and Reproducibility

All analyses were conducted using open-source software. Specifications are described below to enable reproducibility.

## Statistical analysis

If not explicitly reported otherwise, group differences for scalar values were assessed using non-parametric Mann–Whitney U-tests, implemented in scipy (v1.15.3). If applicable, correction for multiple comparisons was performed by using the Benjamini–Hochberg procedure as implemented in statsmodels (v0.14.5).

## Software environment

Motion capture was performed using YOLO11 (yolo11x-pose) from ultralytics (v8.3.187, https://github.com/ultralytics/ultralytics). Feature extraction was carried out using tsfresh (v0.21.0, https://github.com/blue-yonder/tsfresh), tsfel (v0.2.0, https://github.com/fraunhoferportugal/TSFEL), and pycatch22 (v0.4.5, https://github.com/DynamicsAndNeuralSys pycatch22). Experiments with feature-based representations employed gradient boosting xgboost (v1.7.6, https://github.com/dmlc/xgboost). Models using raw time series were implemented with sktime (v0.38.3, https://github.com/sktime/sktime), and dynamic time warping was provided by tslearn (v0.6.4, https://github.com/tslearn-team/tslearn). Logistic regression and k-nearest neighbor classifiers were from scikit-learn (v1.5.2, https://github.com/scikit-learn/scikit-learn). Borderline-SMOTE oversampling was implemented with imbalanced-learn (v0.13.0, https://github.com/scikit-learn-contrib/imbalanced-learn). UMAP dimensionality reduction was implemented using umap-learn (0.5.7, https://github.com/lmcinnes/umap).

## 3 Results

### 3.1 Data

In total, videotaped gait assessments were available from 93 participants originating from three sites. Four video assessments were excluded after visual inspection for failing to meet quality standards, leading to a total of 89 assessments used in this work. Site A data consisted of patients diagnosed with an isolated REM-sleep behavior disorder (iRBD, N=28) as well as healthy controls (HC, N=11); at site B only data from individuals with iRBD (N=30) were assessed; and at site C, only HCs were assessed (N=20). In a subset of 23 individuals with iRBD from site A, results of a diagnostic DaT-Scan were available. The distribution of HCs and iRBDs, as well as respective demographic information and characterizing data, are summarized in Table 2. The investigations into potential shortcut learning, due to video length, age, and sex, did not indicate substantial evidence for such. For more details, see Supplementary Data, Section S3.

**Table 2:** Demographic information and characterizing data of the cohorts. Video recordings of the walking task were acquired at three different sites, denoted as Site A, B, and C, with an imbalanced distribution of the two classes we aimed to distinguish using computer vision-based spatiotemporal motion quantification: patients diagnosed with isolated REM sleep behavior disorder (iRBD) and healthy controls (HC). Demographic and characterizing cohort data are provided per site and for the subset of iRBD from site A in which diagnostic DaT-Scan results were available. Total numbers (N) of iRBD and HC and the female (F)/male (M) split are provided as absolute values. Age, MDS-UPDRS III motor score per class (either iRBD or HC), and the time between DaT-Scan acquisition and the videotaped gait task are provided as mean ± standard deviation (SD). n.a.= not applicable.

| Site | iRBD | HC | Sex | Age | MDS-UPDRS III total score | Timeshift DaT-Scan |
| --- | --- | --- | --- | --- | --- | --- |
|  | N | N | iRBD/HC<br>F:M | iRBD/HC<br>mean±SD | iRBD/HC<br>mean±SD / mean±SD | mean±SD in days |
| A | 28 | 11 | 2:26/2:9 | 69.2 ± 6.1/67.6 ± 5.6 | 9.75 ± 6.21 / 4.81 ± 3.86 | – |
| B | 30 | 0 | 6/24 | 69.7 ± 5.6 | 9.23 ± 5.27 / n.a. | – |
| C | 0 | 20 | 5/15 | 55.0 ± 18.4 | n.a. | – |
| A<sub>DaT-Scan</sub> | 23 | 0 | 0/23 | 69.6 ± 5.5 | 8.56 ± 6.13 / n.a. | −772.17 ± 438.244 |

### 3.2 ML Model Performance

We compared a total of 25 supervised (five model types across five training set configurations, namely site(s) A, A+B, A+C, A+B+C, and B+C) and 10 unsupervised model architectures (five model types across two training set configurations: site B only, and C only) for their ability to discriminate iRBD from HC. Overall, supervised approaches consistently outperformed unsupervised methods in terms of the macro-averaged F_1_-score (see Table 3).

**Table 3:** Classification performance across training-site configurations, for the best-performing supervised and unsupervised ML models, and the human baseline. AUROC, Precision@80% Recall, Balanced Accuracy, and Macro *F*_1_ are reported for discriminating iRBD from HC. Supervised results use the best-performing supervised model type (time series feature extraction with tsfresh combined with XGBoost), evaluated across the training set configurations shown in the left column. Unsupervised results use the best-performing unsupervised model type (tsfresh combined with an isolation forest outlier detection approach), evaluated on training sites B and C. Human baseline performance is included for comparison. Model values are reported as mean *±* standard deviation along with the 95% confidence interval according to 30 repetitions of each experiment with different random initializations; human baseline values are single point estimates. The highest mean AUROC and Precision@80% Recall (supervised) and the highest mean Balanced Accuracy and Macro *F*_1_ (unsupervised) are highlighted in bold font. standard deviation (SD). n.a.= not applicable.

| Train Sites | AUROC | Precision@80%Recall | Balanced Accuracy | Macro $F_1$ |
| --- | --- | --- | --- | --- |
| *Supervised* |  |  |  |  |
| A | $0.615 \pm 0.075$<br>[0.471, 0.743] | $0.760 \pm 0.039$<br>[0.717, 0.838] | $0.557 \pm 0.087$<br>[0.412, 0.691] | $0.553 \pm 0.089$<br>[0.377, 0.681] |
| A + B | $0.638 \pm 0.097$<br>[0.427, 0.776] | $0.773 \pm 0.037$<br>[0.717, 0.834] | $0.575 \pm 0.086$<br>[0.411, 0.706] | $0.570 \pm 0.084$<br>[0.401, 0.694] |
| A + C | $0.672 \pm 0.079$<br>[0.542, 0.799] | $0.813 \pm 0.048$<br>[0.731, 0.885] | $0.645 \pm 0.094$<br>[0.473, 0.774] | $0.647 \pm 0.098$<br>[0.470, 0.775] |
| B + C | $0.482 \pm 0.053$<br>[0.393, 0.582] | $0.722 \pm 0.010$<br>[0.717, 0.757] | $0.472 \pm 0.046$<br>[0.399, 0.583] | $0.427 \pm 0.064$<br>[0.370, 0.586] |
| A + B + C | **$0.739 \pm 0.065$**<br>[0.622, 0.843] | **$0.829 \pm 0.054$**<br>[0.717, 0.900] | $0.672 \pm 0.054$<br>[0.558, 0.752] | $0.676 \pm 0.056$<br>[0.559, 0.764] |
| Human baseline | 0.682 | 0.781 | 0.537 | 0.528 |
| *Unsupervised* |  |  |  |  |
| B | — | — | **$0.609 \pm 0.050$**<br>[0.521, 0.678] | **$0.599 \pm 0.044$**<br>[0.519, 0.661] |
| C | — | — | $0.483 \pm 0.003$<br>[0.482, 0.487] | $0.409 \pm 0.002$<br>[0.409, 0.412] |

Among all evaluated models, the combination of tsfresh features and an XGBoost classifier achieved the highest performance across all training configurations. When trained on the complete cohort (Sites A+B+C), the model peaked with an AUROC of 0.739 *±* 0.065 [0.622, 0.843], a Precision@80%Recall of 0.829 *±* 0.054 [0.717, 0.900], and a macro-averaged F_1_-score of 0.676 *±* 0.056 [0.559, 0.764]. Training on all other training set compilations resulted in reduced performance, with the lowest performance observed when trained exclusively on Site A data, for experiments that included Site A in the training set. Combining training on B+C yielded an even worse performance for the tsfresh+XGBoost model. Increasing the training cohort by adding individuals with iRBD (Sites A+B) yielded only a modest improvement in AUROC (approximately 2%), whereas adding healthy controls (Sites A+C) resulted in a substantially larger performance improvement in AUROC (at least 5%). Performance metrics for the best-performing model for all training configurations are summarized in Table 3 and illustrated in Figure 2. Performance metrics for all other supervised model types are summarized in Supplementary Tables S3; and additional performance measures for the best-performing model, namely the weighted F_1_-score and the area under the precision-recall curve, are reported in Supplementary Table S4.

**Figure 2.**
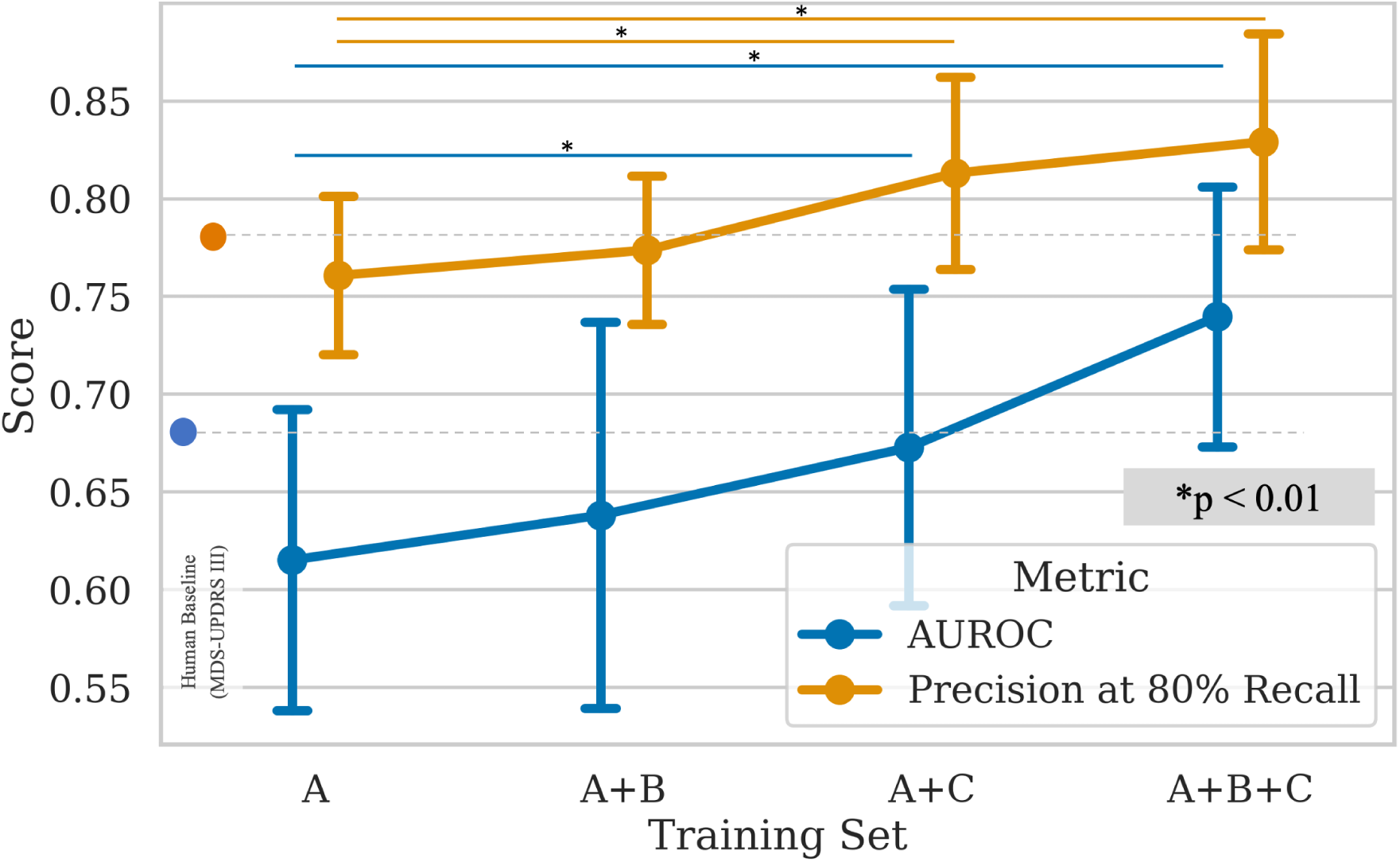
Performance improvement of the best-performing model across different training cohorts. The area under the receiver operator curve (AUROC, shown in blue) and precision at a recall of at least 80% (shown in orange) as performance metrics are visualized across all training set configurations, starting with the training set from site A (iRBD and HC) only and its extension with the iRBD-only (site B) and HC-only (site C) datasets. Mean values and standard deviations are provided. Mean performance by the model using the MDS-UPDRS III score for each metric is given as a human baseline reference, i.e., the blue and orange dots at the very left. The gray dotted line is a helper line to compare model performances to the human baseline. Significant performance improvements between training sets are indicated by asterisks above the respective horizontal bars in blue for AUROC, and orange for Precision@80%Recall, * *p <* 0.01.

Across all unsupervised methods, an isolation forest trained on tsfresh features achieved the highest performance metrics, reaching a macro-averaged F_1_-score of 0.599 *±* 0.044 [0.519, 0.661] and balanced accuracy of 0.609 *±* 0.050 [0.521, 0.678] when trained on Site B (iRBD only) (see Table 3). A comprehensive overview of the performance of all evaluated unsupervised models across all training set configurations is provided in Supplementary Tables S5.

### 3.3 Comparison to Human Baseline

The logistic regression model using the MDS-UPDRS III sum score achieved an AUROC of 0.682 and a Precision@80%Recall of 0.781 with a macro-averaged F_1_-score of 0.528 (see Table 3). Only when trained on data from all sites (A+B+C), the best-performing supervised model architecture, tsfresh+XGBoost, achieved significantly higher average AUROC scores than the human-rated MDS-UPDRS III sum score (p < 0.05). Details of the statistical comparisons between the best-performing ML model and the human baseline are provided in Supplementary Table S6.

### 3.4 Model-guided feature analysis

SHAP values were investigated in the best-performing model architecture, namely tsfresh+XGBoost, on the optimal training set that included all three sites (A+B+C). Across the six body regions, shoulders, elbows, wrists, hips, knees, and ankles, the hips exhibited the highest mean SHAP values averaged across all features derived from time series that included at least one hip marker. Figure 3(I) illustrates the relative magnitude of mean SHAP values for the six aggregated body regions. The overall top three features with the greatest average SHAP values were derived from the following two time series: (1) the difference in the angles at each hip (where the left and right hip are defined as the apex of a respective triangle, each with both ankles forming the base), top1 and top3 features, and (2) the distance between the 2 body markers: left hip and left wrist, top2 feature. The three features with the highest average SHAP values are plotted against each other to further visualize a color-coded class probability overlay calculated from a support vector machine with a sigmoidal kernel, as shown in Figure 3(II). This presentation visually demonstrates how computer-vision-derived features can distinguish between the two classes using a single numerical value.

**Figure 3.**
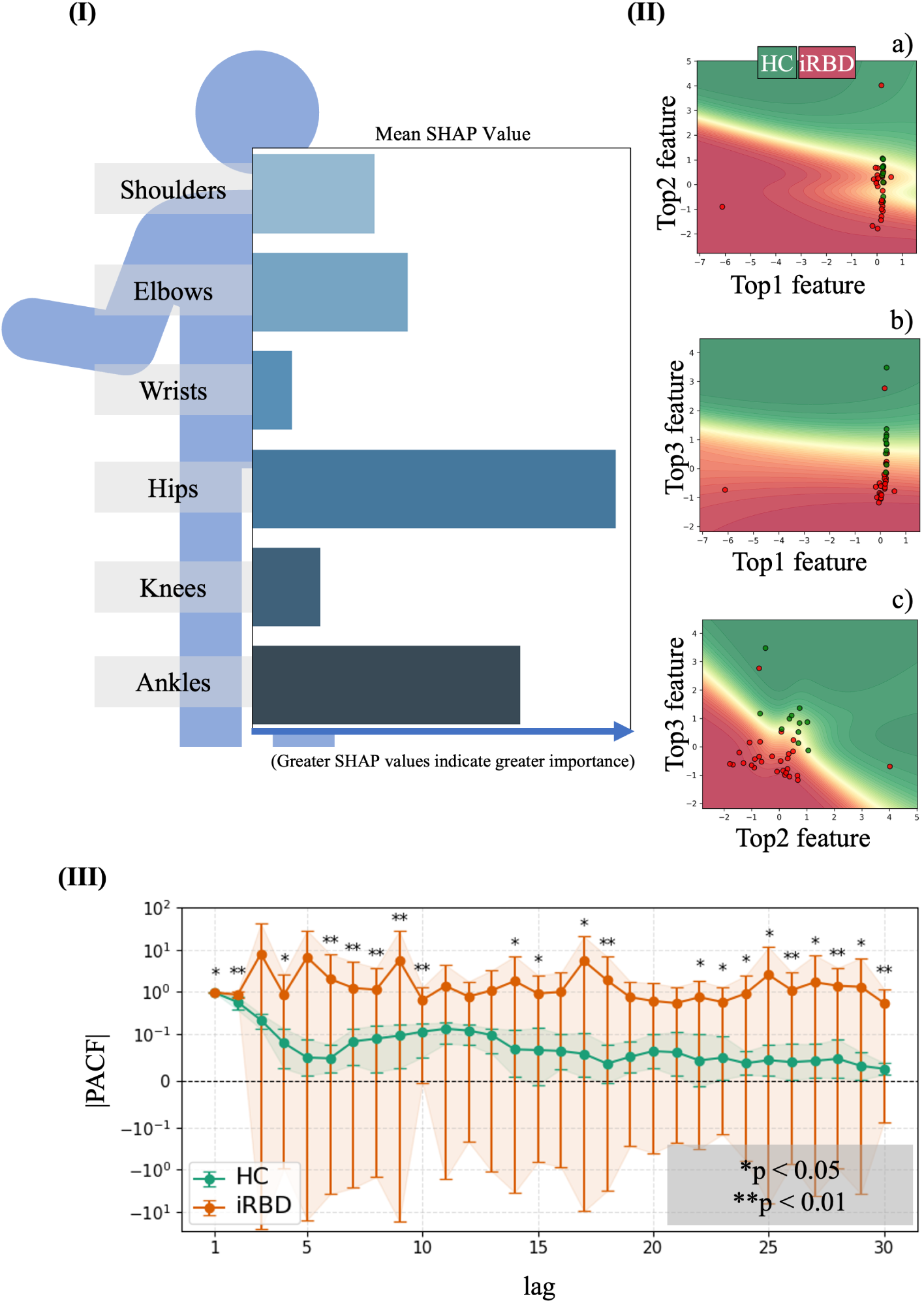
Model-guided analysis of the most influential computer-vision derived digital parameters. Explainable AI with feature attribution analyses could be derived from the best-performing ML model architecture to identify features driving model predictions. The most influential features enabled ranking of the relevant time series and body parts to differentiate iRBD from healthy controls, and informed further exploration of temporal dependencies. (I) Average SHAP values across body positions extracted during the best-performing supervised experiment indicated that the hips had the highest mean SHAP values. (II) The top 3 features for discriminating iRBD from HC are shown pairwise in scatter plots with a class probability overlay calculated from a support vector machine with a sigmoid kernel. The red dots indicate single individuals with iRBD, while green dots depict HC participants. This presentation illustrates visually how computervision-derived features allow for separating the two classes using a single numerical value. Feature details are provided in the Supplementary Data (S8). (III) Partial autocorrelation function (PACF) of the time series describing the difference in the hip angles was evaluated to compare the strength of direct temporal dependencies across lag distances of 1 to 30 between groups. The lags 1-30 are mapped on the x-axis while the y-axis displays the absolute value of the partial autocorrelation function at that specific lag. Mean and standard deviation of the absolute PACF for each lag are provided for iRBD in red and HC in green. The y-axis is displayed with a logarithmic scale for better readability, however resulting in the impression of highly asymmetric error bars. Asterisks indicate lags with significant group differences, \**p_corr_ <* 0.05, \*\**p_corr_ <* 0.01.

Two of the top three features ranked by SHAP values were associated with partial autocorrelation (PAC) at different lags. This motivated a more detailed investigation of the underlying temporal dependencies by analyzing the partial autocorrelation functions (PACF) across lags 1–30 and comparing the temporal structure of the time series underlying the top1 and top3 features between iRBD and HC. The absolute PACF profile, as defined in Section 2.4.2, for the time series underlying the respective SHAP-ranked features (i.e., the difference in hip angles over time), is shown in Figure 3(III). Absolute PACF values were consistently higher in iRBD compared to HC, indicating stronger lag-specific temporal dependencies in the gait-related time series. Differences were statistically significant in more than two-thirds of the investigated lags (Figure 3(III)).

### 3.5 Association with Dopaminergic Imaging

In total, DaT-Scans were available for 23 individuals with iRBD. The availability of neuroimaging measurements varied slightly across striatal regions: caudate (right/left: N = 22/22), anterior putamen (right/left: N = 18/18), posterior putamen (right/left: N = 19/19), and total putamen (right/left: N = 20/21). Demographic and characterization data for the subset of individuals with iRBD from site A with available DaT-Scan measurements are presented in Table 2.

Across all striatal regions, models including only the clinical MDS-UPDRS III score showed the lowest or among the lowest model fit, with R^2^ values consistently below 0.5. Models incorporating the PACF30 embedding generally explained more variance than the MDS-UPDRS III-only models, although the magnitude and uncertainty of the differences varied across regions. The differences in R^2^ between the PACF30 and MDS-UPDRS III models ranged from *−*0.046 in the left putamen to 0.151 in the right posterior putamen, with 95% bootstrap confidence intervals spanning zero across all regions (Supplementary Table S7).

The combination of MDS-UPDRS III and PACF30 consistently yielded higher R^2^ than either predictor set alone. Compared with the MDS-UPDRS III-only model, the combined model increased R^2^ by 0.015 to 0.220 across regions, corresponding to relative increases in explained variance of 7.1% to 186.9%. The largest improvement was observed in the right posterior putamen (ΔR^2^ = 0.220, 95% CI [0.001, 0.568]), whereas the smallest was observed in the left caudate (ΔR^2^ = 0.015, 95% CI [0.000, 0.271]). The corresponding differences between models are summarized in Supplementary Table S7.

The highest model fit was observed for the combined MDS-UPDRS III + PACF30 model in the left posterior putamen, with R^2^ = 0.479 (95% CI [0.291, 0.858]). Although this represented the best-performing model among the MDS-UPDRS III-only, PACF30-only, and combined models, none of these models exceeded an R^2^ of 0.5 in any region, indicating that even the best-performing model explained less than half of the variance in DaT-Scan outcomes. The combined model also consistently showed higher R^2^ than the PACF30-only model, with ΔR^2^ values ranging from 0.019 in the left anterior putamen to 0.222 in the left posterior putamen (Supplementary Table S7). The corresponding AIC and AIC*_c_* values were generally lower for models incorporating PACF30, either alone or in combination with MDS-UPDRS III, compared with the MDS-UPDRS III-only model, indicating improved model fit after accounting for model complexity. The R^2^ values per model and the respective increase, both absolute and relative, are shown in Figure 4.

**Figure 4.**
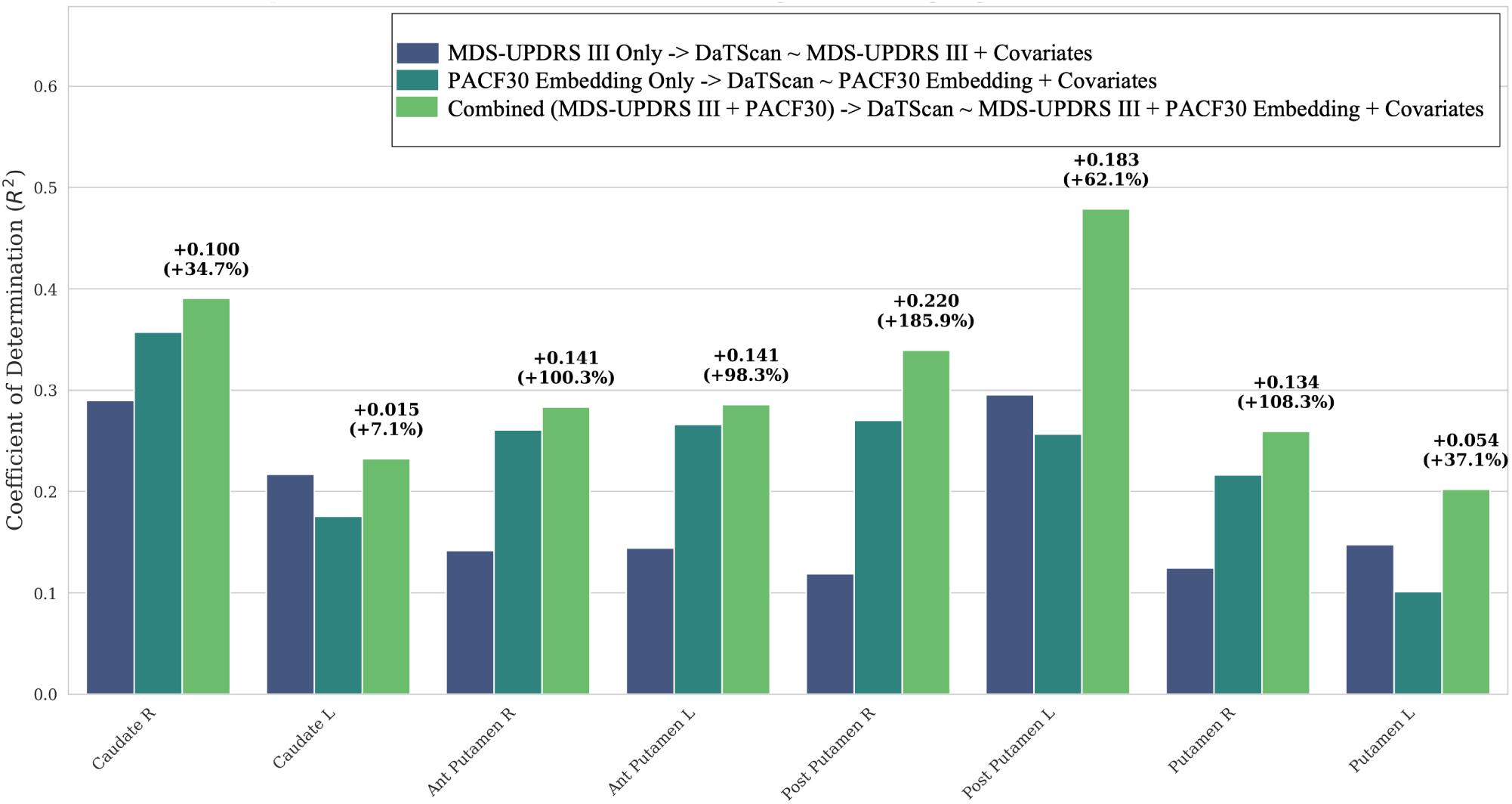
Association of clinical and computer-vision-derived parameters with dopaminergic imaging across striatal regions. Striatal regions are shown along the x-axis, with bar heights representing the mean coefficient of determination (*R*^2^) obtained from bootstrap regression models predicting regional binding ratios (DaT-Scan z-scores) from the MDS-UPDRS III score, the model-informed computer-vision-derived PACF30 embedding, or their combination (MDS-UPDRS III + PACF30). The percentage above the rightmost bar in each region indicates the relative gain in *R*^2^ from the MDS-UPDRS III-only model to the combined MDS-UPDRS III + PACF30 model. R = right, L = left, Ant = anterior, Post = posterior.

In contrast, models based on the TOP30 embedding generally showed comparable or lower model fit than models incorporating the PACF30 embedding. Comparable performance between TOP30 and PACF30 models was observed in only two regions. Detailed results for all models, including R^2^, AIC, and AIC*_c_*, are provided in Supplementary Tables S9, S10, and S11.

## 4 Discussion

Multi-site video data of a standard walking task acquired with a single monocular RGB camera from patients diagnosed with an isolated REM sleep behavior disorder (iRBD) and healthy controls (HC) were used to evaluate the potential of computer-vision-derived parameters to identify group differences in gait patterns and inform in-depth analyses of the most influential kinematic features, including correlations with dopaminergic imaging. Kinematic time series were extracted using a standard motion capture technique. Ten different supervised and unsupervised machine learning (ML) models were trained to distinguish between HC and iRBD across various training set configurations, ranging from fully confounded single-site data to the entire cohort data from all three sites, while still handling highly imbalanced samples across sites. The overall best-performing model was a supervised ML architecture that used time series features, extracted with the tsfresh library, rather than the raw time series, and XGBoost, a gradient-boosted decision tree algorithm for supervised classification and regression. Model performance increased across further enrichment of training data, peaking when trained on the entire cohort with data from all three sites (Sites A+B+C), with an average AUROC of 0.739. In contrast, fitting a model on the best-performing clinical reference, MDS-UPDRS III score, yielded an AUROC of 0.682, which was considered the human baseline performance in this study. Thus, our analyses indicate that expanding the dataset with additional HC and iRBD samples can substantially improve ML model performance, even in the presence of class imbalance.

Notably, neither training the tsfresh+XGBoost model solely on the baseline training dataset from the single site containing both classes (Site A), nor enriching the training dataset alone with additional single-class iRBD or HC data (Sites A+B/C), enabled the model to exceed the clinical baseline performance (AUROC: 0.615, 0.638, and 0.672, respectively, compared to 0.682 for the clinical baseline).

ML approaches generally benefit from larger training datasets, and high data quantity alone cannot compensate for limited data quality [75]. Although we applied several strategies to mitigate site effects and reduce the risk of shortcut learning, it remains unclear to what extent these approaches generalize to more heterogeneous datasets and alternative input modalities, such as hand-function assessments or sensor-derived data. A key advantage of video-based input was the ability to verify task execution directly, reducing the reliance on outlier detection methods that must distinguish acquisition errors from valid observations within the expected data distribution.

Unsurprisingly, the performance of unsupervised approaches was inferior to that of supervised ML models, in terms of macro-averaged F_1_-score. But although unsupervised methods did not achieve human baseline performance, they attained balanced accuracy scores above chance, indicating that the models made non-random distinctions between individuals with iRBD and HC. These performance results indicate the presence of measurable alterations in gait behavior in individuals with iRBD compared to HCs using digital assessments even derived from the very simple set-up of a back-and-forth standard walking task captured with a single monocular RGB camera.

The walking task conducted in this study consisted of normal-speed walking without additional cognitive or motor challenges. Previous studies have reported gait alterations in iRBD primarily during more demanding walking conditions, such as fast-paced walking or dual-task paradigms, while differences during standard walking were less consistently observed [29]. However, the study by *Martens et al.* [29] relied on a pressure-sensor carpet, suggesting that differences in measurement modality and feature extraction may influence the sensitivity for detecting subtle gait alterations in iRBD. One potential advantage of video-based assessment is the simultaneous capture of multi-segment body kinematics, enabling the characterization of coordinated movement patterns beyond predefined gait parameters. This interpretation remains speculative and should be investigated in future studies directly comparing video-based and sensor-based approaches within the same cohort.

Furthermore, our findings are consistent with previous reports using quantitative motor assessments, that demonstrated subtle gait and movement alterations in individuals with iRBD despite the absence of clinically manifest parkinsonism [18, 28–36]. Previous sensor-based studies have described alterations in gait symmetry, stride characteristics, movement amplitude, and arm swing in iRBD, suggesting that early motor abnormalities are not restricted to clinically observable signs but can already be detected through quantitative measurements [30–32, 37]. Our results extend these observations by demonstrating that subtle motor differences can be captured from a minimalistic video setup without dedicated wearable sensors or specialized laboratory infrastructure.

Importantly, markerless motion capture enables extraction of high-dimensional kinematic time series that describe coordinated body movement over time. Such data-driven representations may allow the identification of complex movement characteristics that are difficult to capture using predefined sensor-derived parameters alone. Therefore, simple video-based assessments may represent a promising approach for detecting subtle motor alterations associated with prodromal neurodegeneration. However, future studies directly comparing video-based and sensor-based approaches within the same individuals will be required to determine whether video-derived measures provide complementary information or greater sensitivity to early motor changes.

Next, we aimed to use the data-driven ML model architecture as guidance for further in-depth analyses. For these model-guided additional downstream analyses, we used an explainable AI approach using SHAP values, enabling ranking of the most influential parameters to distinguish motion patterns of iRBD from HC. The hips were identified as the most influential body parts, particularly time series including the angles from one hip, with both ankles as the triangle basis. At the feature level, particular partial autocorrelation (PAC) at different lags was repeatedly identified as an important feature. Further comprehensive investigations of PAC across a broad range of lag intervals confirmed a consistent pattern of higher values in iRBD than in HC, with significance in most lags. Moreover, embeddings of these PAC functions to achieve a scalar representation exhibited higher correlations with DaT-Scan-derived regional z-scores than clinical scale-only embedding or embeddings of the overall top 30 ranked features. Correlating the regional z-scores of dopaminergic uptake with a combination of clinical scales, MDS-UPDRS III score, and PAC function embeddings resulted in R^2^-score increases of at least 7.1%, and up to 185.9%. Even though such overall data-driven approaches should not generally be limited by their interpretability in terms of known clinical observations, it remains notable that the model-guided analyses revealed PAC functions as influential and relevant parameters in iRBD vs. HC. Higher absolute PAC values indicate a greater linear dependence related to that specific lag [76]. Thus, the consistently higher absolute PAC values in iRBD across a broad range of lags might be interpreted as an overall more predictable temporal structure of the signal. While speculative, this increased temporal dependence may reflect an early rigidity-like motor phenotype in iRBD, in which movements become more stereotyped and less flexible. This finding aligns with prior studies using wearable motor sensors, which reported increased trunk rigidity and reduced flexibility in individuals with iRBD [30].

Even with the limited sample size and the temporal delay of diagnostic DaT-Scan dopaminergic imaging, our analysis revealed consistently higher explanatory power of model-guided digital gait feature embedding, both alone and in combination with the clinical MDS-UPDRS III scale, compared with the clinical scale alone. Although no causal conclusions can be drawn, this might indicate that digital motor biomarkers may provide at least complementary and possibly more sensitive proxies for underlying nigrostriatal degeneration in iRBD. Broadly related yet interestingly, motor behavior has been correlated with cognitive performance when assessed behavior using a multi-camera motion capture system [77]. Such findings, alongside those of the presented work, strongly suggest further research into the interplay among neurodegenerative pathology markers, cognition and movement markers, and the extent to which one determines the other. Additionally, future studies should examine how including motor reserve as a covariate influences model prediction. In general, when relating the findings of this study to the wider field of iRBD and movement disorders research, certain achievements are notable. First, the multi-site approach has successfully demonstrated feasibility and the advantage of enriching training data, even with unbalanced data, laying the foundation for future multi-site movement cohort studies to consider the inclusion of video gait data. Notably, even though a very simple and easy-to-implement video setup with a single monocular RGB camera, like in most smartphones, was utilized in this study, our results demonstrated that video-derived motion capturing remained robust across sites. This stability highlights the suitability of the chosen approach for larger-scale studies, offering the inclusion of underrepresented areas with limited equipment. Second, the SHAP analysis in this work demonstrates that integrating explainable AI into modeling approaches provides a deeper understanding of the underlying patterns by supporting a ranking of interpretations of the findings and informing further downstream analyses in a model-guided fashion focusing on parameters being identified as highly influential.

With regard to the potential for clinical translation, the best-performing model achieved a precision of 82.9% while maintaining a recall of at least 80%. This qualifies the presented approach as a potential low-cost early screening tool for iRBD that can be easily integrated into routine visits beyond specialized movement disorder clinics. Unlike the MDS-UPDRS III, which requires trained clinicians and is subject to inter-rater variability, this video-based digital assessment provides objective, standardized measurements that can be deployed at scale in routine clinical practice. In general, the two aspects, high accuracy and interpretability, may qualify the presented work as a foundation for future efforts towards video-based motion assessments as a decision support tool.

Even though several strategies were implemented to mitigate the risk of site-related shortcut learning and biased performance evaluation, e.g., class-imbalance correction and class-aware cross-site harmonization, the class imbalance across sites remains one of the major limitations of the present study on retrospective data sets. While two sites comprised one-class data only, even the site that holds both classes, iRBD and HC, was substantially imbalanced towards the iRBD class. Given this imbalance (28 iRBD vs. 11 HC), a trivial classifier that predicts iRBD for every sample would already achieve a precision of 71.8%, which should be considered when interpreting the absolute precision of 82.9% achieved by the best-performing model. As a result, this limits the selection of suitable samples for a fair test set. A further limitation is that class-aware cross-site harmonization cannot be applied prospectively, as it requires knowledge of sample class labels that would not be available for new, unseen cases in routine clinical use. Furthermore, the class imbalance on the test site makes certain commonly used performance metrics, such as the area under the precision-recall curve or the non-balanced accuracy, incapable of accurately assessing the performance. Further recruitment at additional sites is necessary to identify or rule out site effects. Relatedly, the test set used for evaluation was small (n = 39), and the presented findings have not yet been validated on an independent external dataset; both factors limit the precision of the reported performance estimates and the generalizability of the results. Another limitation arises when considering the feasibility of translating our approach into clinical practice. In this study, we relied on manually quality-controlling each sample. Even though time-consuming, this is in turn an advantage of video data compared to, e.g., sensor-derived digital measures. Visual inspection provides precise control in determining whether an outlier value is due to factors such as incorrect task execution, unexpected interruptions, or participant mislabeling, or represents a genuine instance of extreme movement behavior. However, especially in larger multi-center scenarios, this will become increasingly challenging. Nevertheless, once automated quality control is implemented, video-based assessment will offer substantial scalability advantages over repeated UPDRS-III ratings, which are time-intensive and require specialized expertise at every assessment point. Thus, future developments will prioritize automated quality control to fully realize the scalability and standardization benefits of digital gait assessment over conventional clinical rating scales. Ideally, such methods would not only flag obvious outliers in the final analyses after data collection, but also provide immediate quality feedback right after acquisition, to ensure a consistent standard across sites and reduce variability in data quality.

Furthermore, we want to acknowledge that there is no default recommendation for ML model architecture design. In the presented use case, model performance on the classification task of distinguishing iRBD from HC was volatile and, in most cases, inferior to the clinical MDS-UPDRS III sum score, considered the human baseline. Combinations of the best-performing model classifier XGBoost with other feature extractors, namely tsfel and catch22, were less accurate than its combination with tsfresh. As tsfresh includes the largest feature library, one possible interpretation is that the reduced feature sets of the alternative tools simply do not extract the features that encode the underlying movement patterns to distinguish iRBD and HC. Regarding the ML model architecture (Mini-)ROCKET, which operates directly on the raw time series without feature extraction, the limited sample size could be a possible cause of the insufficient performance. Finally, the combination of dynamic time warping (DTW) and a k-nearest neighbor classifier ranked second after the tsfresh+XGBoost model. However, it neither reached human-level performance nor showed the same improvement as increasing training data. This may indicate that DTW-based similarity measures do not adequately capture the disease-specific temporal dynamics needed to distinguish iRBD and healthy controls.

Taken together, these results highlight the importance of systematically evaluating different machine learning architectures for a given research question, particularly when working with small sample sizes and emerging data modalities such as video-based motion capture. Clinical assessment scales, such as the MDS-UPDRS III, were specifically designed to capture manifestations of Parkinson’s disease and therefore incorporate domain knowledge about the underlying pathology. Similarly, machine learning research should not assume that a single model architecture is universally optimal; instead, it should investigate and develop architectures best suited to capturing disease-specific characteristics in the available data. Given these limitations, future investigations should evaluate the best-performing model, namely tsfresh+XGBoost, on a class-balanced external dataset to assess its performance and ability to generalize to unseen data captured in a different setting. In the same manner, any future research shall investigate whether PAC functions serve as suitable biomarkers that exhibit similar behavior, with higher absolute values for iRBD across other datasets. The cautiously framed discussion of the observed associations between PAC functions and markers of neurodegeneration, quantified using regional z-scores of dopaminergic uptake, highlights the need for further investigation in larger and more comprehensive cohorts to confirm the findings. If confirmed, such digital readouts could represent a practical tool for monitoring disease progression and for use in clinical trials, reducing reliance on time-intensive clinical ratings while preserving sensitivity to dopaminergic dysfunction.

Finally, although feature-based methods seem promising for the given iRBD detection task, other models operating directly on the raw time series warrant future investigation. This shall include neural network architectures such as LSTMs and CNNs, as well as the use of pre-trained models for more accurate iRBD detection based on videotaped walking tasks. This, however, requires more computational effort and larger datasets and was considered unsuitable and out of scope for this work.

The presented models were thoroughly evaluated, and a candidate was isolated that holds potential for translation into clinical practice. The evaluation scheme enables a realistic estimate of expected performance while presenting a multi-site framework that improves the human baseline by a meaningful margin. The presented work uses three datasets and evaluates various scenarios for leveraging this data. This includes both supervised and unsupervised experiments, as well as all possible combinations of training sets. This comprehensive methodology allows a wide overview of how this type of data can be utilized for the underlying task of iRBD detection. However, a central aspect of the study goes beyond the pure classification task, but rather aims to leverage explainable AI to identify the most influential parameters and body parts, and thus, in a model-guided, data-driven manner, inform further in-depth analyses focusing on the preselection of high-impact parameters. Summarizing the strengths of this work, this multi-site, multi-paradigm machine learning study achieved promising performance, identified digital movement biomarkers (PAC functions) that provided interpretable insights into the movement behaviors distinguishing iRBD from HC, and ultimately generated a preliminary hypothesis regarding a potential association between digital movement markers and neurodegenerative processes assessed by dopaminergic imaging.

Beyond the future investigations outlined above, an important next step is the continued recruitment of participants at the existing study sites, as well as the inclusion of additional cohorts to further assess the generalizability of the proposed approach and to support longitudinal follow-up. Moreover, continued efforts towards standardized acquisition protocols and automated quality control procedures will be essential to ensure robust and reproducible movement analysis across sites. Furthermore, future studies should examine sex-specific gait patterns in Parkinson’s disease, given evidence of sex differences in motor presentation and progression [78, 79]. Such developments may facilitate the future evaluation of markerless motion capture as a digital assessment tool for iRBD and related movement disorders.

Collectively, our findings imply that digitally derived gait measures can capture aspects of motor dysfunction not fully captured by conventional clinical rating scales. Unlike the MDS-UPDRS III, which relies on expert assessment and is hence prone to subjectivity, digital gait readouts are objective and rater-independent, offering a scalable and standardized alternative.

### 4.1 Conclusion

The present study demonstrates that movement characteristics extracted from a simple walking task video-recorded with a single standard monocular RGB camera, using markerless motion capture, provide clinically relevant information for distinguishing individuals with iRBD from healthy controls. However, the primary contribution of this study extends beyond classification performance. By leveraging explainable AI, we identify the most influential features and body parts, and these model-derived insights guided subsequent in-depth, data-driven analyses. The identified parameters of the partial autocorrelation of the hip angle motor time series exhibited iRBD-characteristic patterns and correlated with dopaminergic imaging. Taken together, our findings support further investigation of video-based markerless motion capture as an objective and scalable approach for quantitative motor assessment in iRBD. Larger prospective studies are needed to determine its clinical utility and potential role in future screening or decision-support workflows.

## 5 Author Contributions

AO, GRF, EK, and MS conceptualized the studies on sites A and B and provided resources for data collection. AO, SR, KKu, CD, AS, KKo contributed to recruitment, data collection, and clinical scoring for sites A and B. MGE, JF, and KF contributed to the data collection on site C. MS, JF, and PW conceptualized this project. PW created the code, performed the analysis, and wrote the first draft of the paper. All authors revised the manuscript and approved the final version for submission.

## Data Availability

Data will be made available upon reasonable request. The data from site C is available in the form of videotaped assessments alongside
a table providing the clinical ratings, as well as other characterizing information such as age. All requests concerning site C shall
be addressed to the corresponding author, Philipp Wegner. The data from sites A+B are not available
for sharing. All requests concerning sites A+B shall be addressed to Michael Sommerauer.

## 6 Acknowledgments

PW received funding from the iBehave Network, sponsored by the Ministry of Culture and Science of the State of North-Rhine-Westphalia. The setup and data collection on site A were supported by the Koeln Fortune Program / Faculty of Medicine, University of Cologne (grant no. 329/2021, AO), the “Novartis-Stiftung für therapeutische Forschung” (AO). M.S. received funding from the program “Netzwerke 2021”, an initiative of the Ministry of Culture and Science of the State of Northrhine Westphalia, the Federal Ministry of Research, Technology and Space (BMFTR) under the funding code (FKZ) 01EO2107 and under the umbrella of the Partnership Fostering a European Research Area for Health (ERA4Health) (GA N° 101095426 of the EU Horizon Europe Research and Innovation Programme), and the European Research Council (ID 10116958).

## 8 Data Availability

Data will be made available upon reasonable request. The data from site C is available in the form of videotaped assessments alongside a table providing the clinical ratings, as well as other characterizing information such as age. All requests concerning site C shall be addressed to the corresponding author, Philipp Wegner. The data from sites A+B are not available for sharing. All requests concerning sites A+B shall be addressed to Michael Sommerauer.

## Supplementary Material

### S1 List of all time series

**Table S1:** Complete list of all time series considered in this work and how they are derived. List of all 14 time series derived from the 12 body markers (joint position of shoulder, elbow, wrist, hip, knee and ankle, each left + right) initially extracted from the motion-captured walking task.

| Time series name | Description |
| --- | --- |
| dist_left_wrist-left_hip | Distance from left hip to left wrist |
| dist_right_wrist-right_hip | Distance from right hip to right wrist |
| dist_left_elbow-right_elbow | Distance between left and right elbow |
| dist_left_wrist-right_wrist | Distance between left and right wrists |
| dist_left_knee-right_knee | Distance between left and right knees |
| dist_left_ankle-right_ankle | Distance between left and right ankle |
| angle_right_shoulder-(right_wrist-right_hip) | Angle at right shoulder, defined as the apex of a triangle formed by the right wrist and the right hip as the base |
| angle_left_shoulder-(left_wrist-left_hip) | Angle at left shoulder, defined as the apex of a triangle formed by the left wrist and the left hip as the base |
| angle_left_hip-(left_ankle-right_ankle) | Angle at left hip, defined as the apex of a triangle formed by both ankles as the base |
| angle_right_hip-(right_ankle-left_ankle) | Angle at right hip, defined as the apex of a triangle formed by both ankles as the base |
| area_shoulders-hips | Area of the rectangle formed by both shoulders and both hips |
| area_hips-ankles | Area of the rectangle formed by both hips and both ankles |
| diff_shoulders | Difference in the angles between the left and right shoulder triangles (angle_left_shoulder-(left_wrist-left_hip and angle_right_shoulder-(right_wrist-right_hip)) |
| diff_hips | Difference in angles between left and right hip triangles (angle_left_hip-(left_ankle-right_ankle) and angle_right_hip-(right_ankle-left_ankle)) |

### S2 Class-aware cross-site harmonization details

To mitigate site-induced distributional shift, we align features from each single-class site B or C to the distribution of site *A prior to training*, leaving the training data unchanged. The following paragraph illustrates this process as an example for site B. Let **X***_A_ ∈* R*^nA×d^* be the training feature matrix and **X***_B_ ∈* R*^nB×d^* the external feature matrix, where d is the feature dimensionality.

#### S2.0.1 Class-Conditional Mean and Variance Alignment

For each external site whose samples belong to class c, we restrict the reference statistics to the subset of training samples sharing that class:

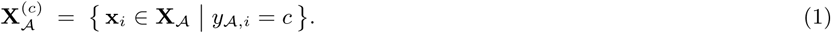

This prevents class-imbalance artefacts from contaminating the alignment signal. We then standardise each external feature to zero mean and unit variance and rescale it to match the first two moments of the corresponding training distribution. Formally, for each feature j = 1,…,d:

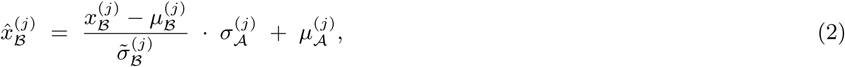

where *μ_A_^(j)^* are the feature-wise mean and standard deviation computed over **X***_A_*^(*c*)^, and µ*_B_*^(*j*)^, σ*_B_*^(*j*)^ are the corresponding statistics of **X***_B_*. The term σ̃*_B_^(j)^*=max (σ*_B_^(j)^*,ε) prevents division by zero for constant features. The full procedure is given in Algorithm 0.

### S2.1 Class-aware alignment

#### Require

Training data (**X***_A_*, **y***_A_*) from home site *A*; external site data (**X***_B_*, **y***_B_*)

#### Ensure

Aligned external features X B

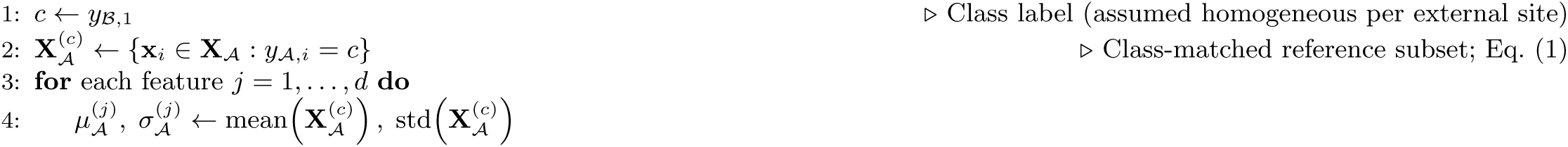

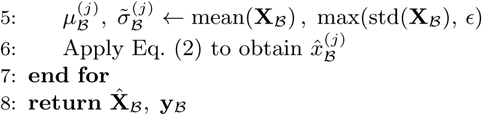

**Figure S1:**
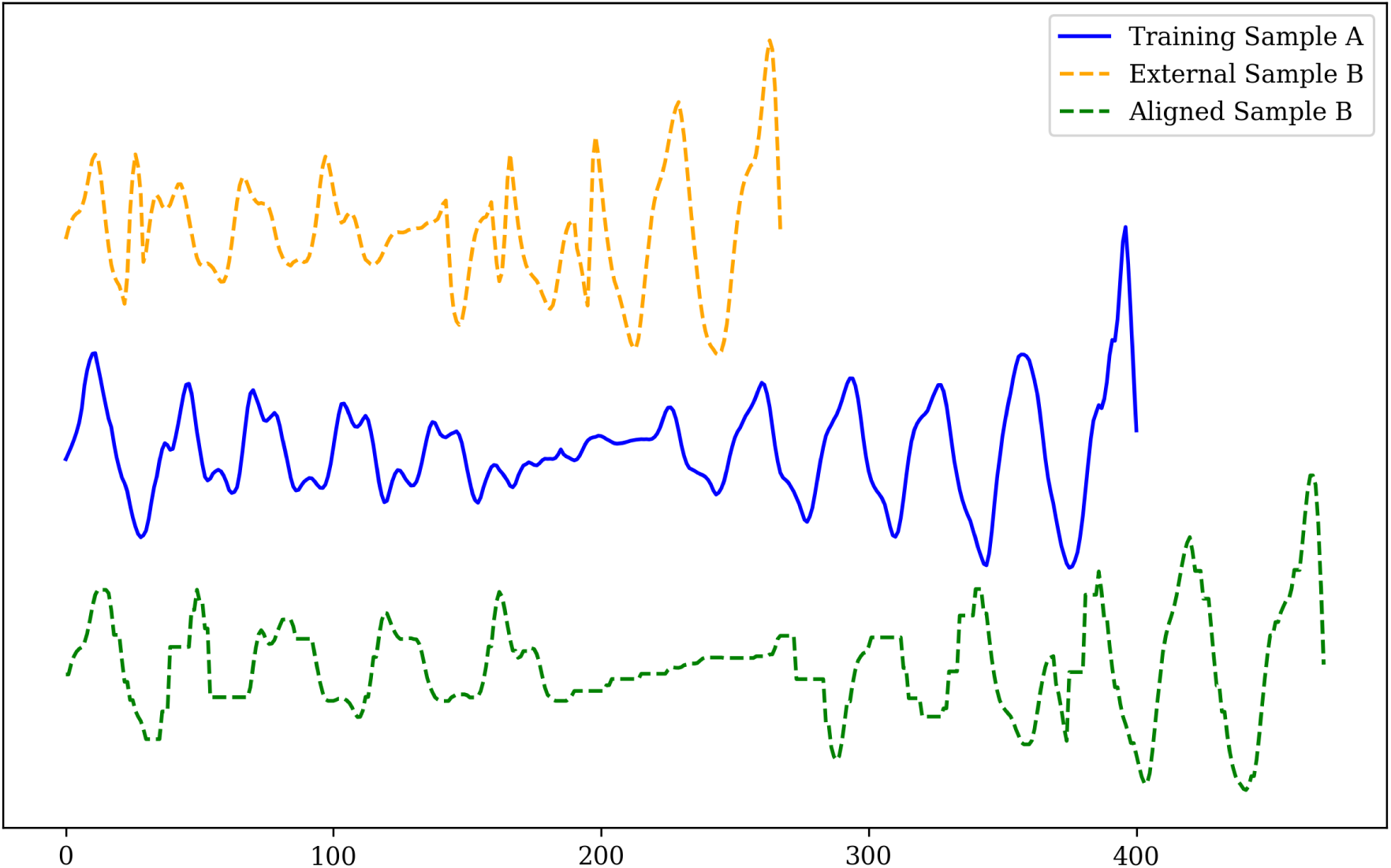
Example of dynamic time warping (DTW) alignment. The upper time series presents a sample from an external site, B or C. The center time series shows its closest, in terms of DTW distance, sample in A. The lower time series presents the external time series aligned to its corresponding sample in A.

### S3 Shortcut Learning

Before undertaking the actual predictive modeling, an investigation was conducted to identify potential confounders that could lead to shortcut learning. This analysis was conducted on site A. Logistic regression analysis was performed to assess whether age, sex, and video length confound the relationship with the class variable (RBD or HC); Group *∼* age+sex+video length. None of these variables showed a significant association with the class (sex: p = 0.229; age: p = 0.309; video length: p = 0.921). The overall model was not significant (LLR p = 0.556), indicating that age, sex, and video length are unlikely to act as confounders in this context, and shortcut learning via these variables is doubtful.

### S4 Hyperparameters

**Figure S2:**
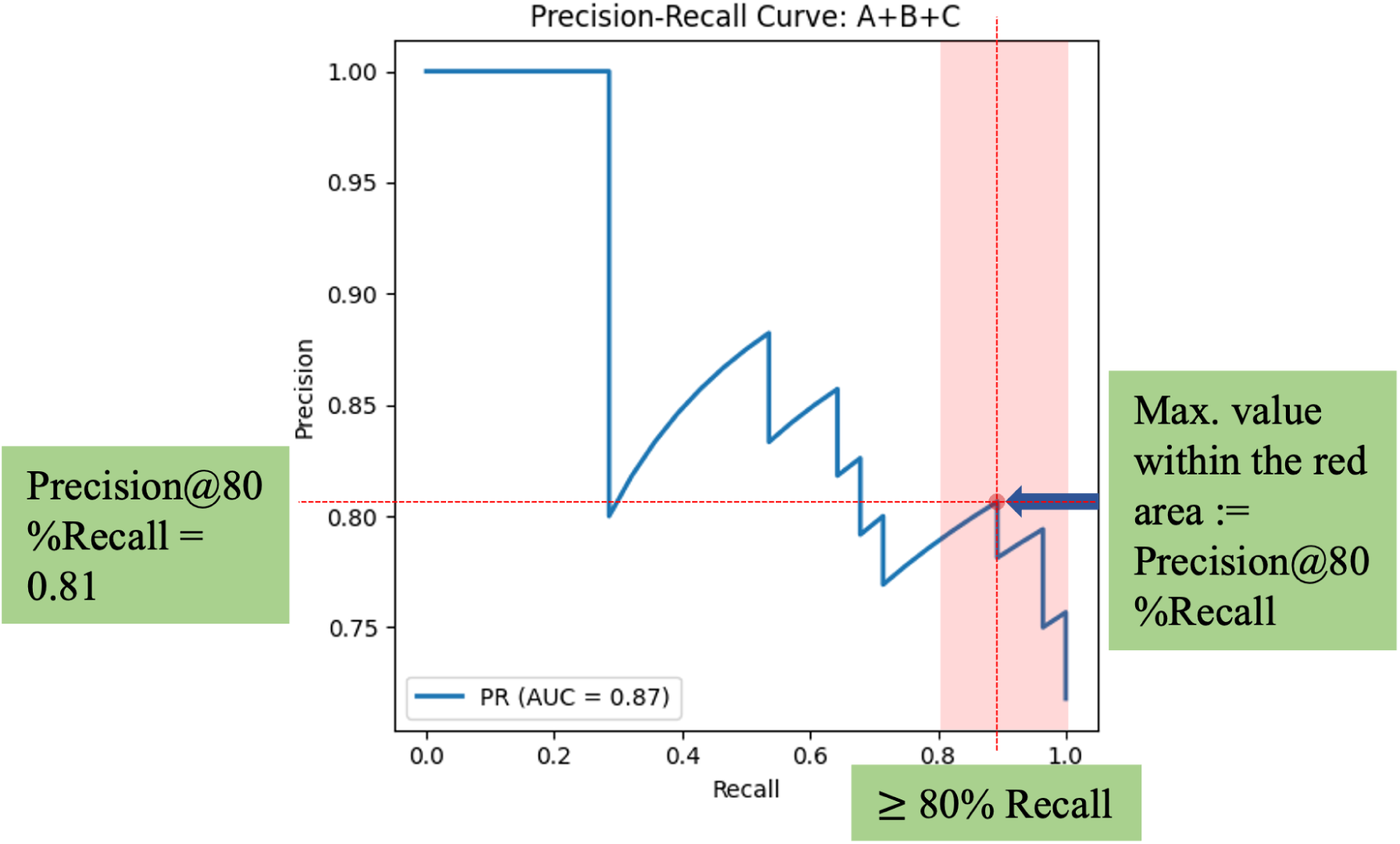
Precision at constrained recall. Precision values are reported at a recall level of at least 80%, defined as the highest precision attainable on the precision–recall curve while maintaining recall above the specified threshold. This metric characterizes model performance under high-sensitivity operating conditions, reflecting its suitability for applications in which missing positive cases is costly.

**Table S2:**
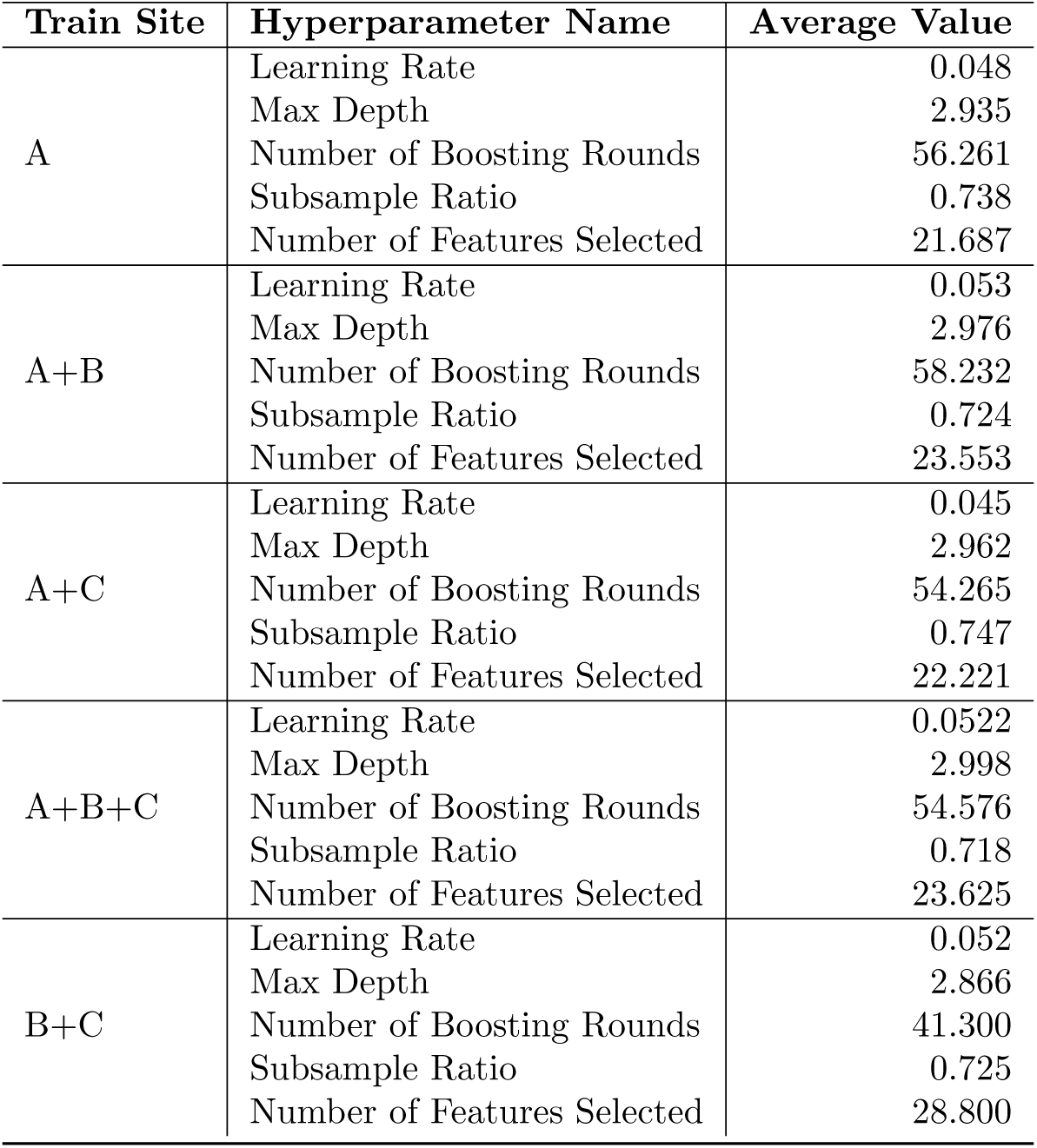
Hyperparameter averages across the best-performing supervised experiment. Hyperparameters were tuned for each training fold within the leave-one-out cross-validation. The values reported here are averages across all folds.

### S5 ML model evaluation metrics for supervised and unsupervised model architectures

**Table S3:** Classification performance of the supervised model types. Model performance for discriminating iRBD from HC is reported as area under the receiver operating characteristic curve (AUROC), precision at 80% recall for each training set configuration, balanced accuracy, and macro-averaged *F*_1_-score (Macro *F*_1_). Values are reported as the mean and standard deviation alongside the 95% confidence interval according to the 30 repetitions of each experiment with different random initializations.

| Train Sites | AUROC | Precision@80%Recall | Balanaced Accuracy | Macro $F_1$ |
| --- | --- | --- | --- | --- |
| **tsfresh+XGBoost (S3) - see main document** |  |  |  |  |
| **tsfel+XGBoost (S4)** |  |  |  |  |
| B+C | $0.502 \pm 0.057$<br>[0.416, 0.622] | $0.725 \pm 0.016$<br>[0.718, 0.774] | $0.485 \pm 0.043$<br>[0.406, 0.581] | $0.454 \pm 0.052$<br>[0.384, 0.566] |
| A | $0.631 \pm 0.067$<br>[0.503, 0.741] | $0.759 \pm 0.036$<br>[0.718, 0.830] | $0.554 \pm 0.066$<br>[0.430, 0.661] | $0.554 \pm 0.069$<br>[0.430, 0.670] |
| A + B | $0.532 \pm 0.063$<br>[0.421, 0.672] | $0.749 \pm 0.025$<br>[0.718, 0.823] | $0.532 \pm 0.050$<br>[0.461, 0.643] | $0.530 \pm 0.052$<br>[0.458, 0.648] |
| A + C | $0.613 \pm 0.052$<br>[0.542, 0.714] | $0.751 \pm 0.024$<br>[0.718, 0.800] | $0.523 \pm 0.047$<br>[0.440, 0.613] | $0.521 \pm 0.050$<br>[0.428, 0.618] |
| A + B + C | $0.506 \pm 0.064$<br>[0.388, 0.612] | $0.725 \pm 0.011$<br>[0.718, 0.759] | $0.442 \pm 0.053$<br>[0.339, 0.540] | $0.428 \pm 0.057$<br>[0.328, 0.534] |
| **catch22+XGBoost (S5)** |  |  |  |  |
| B+C | $0.479 \pm 0.075$<br>[0.321, 0.613] | $0.718 \pm 0.000$<br>[0.718, 0.718] | $0.463 \pm 0.018$<br>[0.438, 0.485] | $0.401 \pm 0.013$<br>[0.391, 0.424] |
| A | $0.465 \pm 0.067$<br>[0.339, 0.581] | $0.738 \pm 0.016$<br>[0.718, 0.772] | $0.465 \pm 0.059$<br>[0.355, 0.587] | $0.462 \pm 0.061$<br>[0.349, 0.588] |
| A + B | $0.536 \pm 0.051$<br>[0.461, 0.642] | $0.727 \pm 0.012$<br>[0.718, 0.755] | $0.497 \pm 0.070$<br>[0.407, 0.693] | $0.494 \pm 0.065$<br>[0.410, 0.670] |
| A + C | $0.365 \pm 0.061$<br>[0.243, 0.479] | $0.730 \pm 0.011$<br>[0.718, 0.750] | $0.447 \pm 0.049$<br>[0.357, 0.555] | $0.431 \pm 0.056$<br>[0.339, 0.556] |
| A + B + C | $0.540 \pm 0.048$<br>[0.453, 0.610] | $0.731 \pm 0.013$<br>[0.718, 0.767] | $0.449 \pm 0.044$<br>[0.382, 0.534] | $0.437 \pm 0.050$<br>[0.358, 0.534] |
| **DTW+kNN (S1)** |  |  |  |  |
| B+C | $0.482 \pm 0.000$<br>[0.482, 0.482] | $0.718 \pm 0.000$<br>[0.718, 0.718] | $0.482 \pm 0.000$<br>[0.482, 0.482] | $0.409 \pm 0.000$<br>[0.409, 0.409] |
| A | $0.660 \pm 0.004$<br>[0.656, 0.669] | $0.806 \pm 0.000$<br>[0.806, 0.806] | $0.648 \pm 0.000$<br>[0.648, 0.648] | $0.639 \pm 0.000$<br>[0.639, 0.639] |
| A + B | $0.667 \pm 0.002$<br>[0.662, 0.671] | $0.806 \pm 0.000$<br>[0.806, 0.806] | $0.648 \pm 0.000$<br>[0.648, 0.648] | $0.639 \pm 0.000$<br>[0.639, 0.639] |
| A + C | $0.641 \pm 0.000$<br>[0.641, 0.641] | $0.800 \pm 0.000$<br>[0.800, 0.800] | $0.630 \pm 0.000$<br>[0.630, 0.630] | $0.617 \pm 0.000$<br>[0.617, 0.617] |
| A + B + C | $0.640 \pm 0.003$<br>[0.636, 0.643] | $0.800 \pm 0.000$<br>[0.800, 0.800] | $0.630 \pm 0.000$<br>[0.630, 0.630] | $0.617 \pm 0.000$<br>[0.617, 0.617] |
| **(Mini-)ROCKET (S2)** |  |  |  |  |
| B+C | $0.547 \pm 0.004$<br>[0.537, 0.553] | $0.718 \pm 0.000$<br>[0.718, 0.718] | $0.464 \pm 0.000$<br>[0.464, 0.464] | $0.400 \pm 0.000$<br>[0.400, 0.400] |
| A | $0.575 \pm 0.035$<br>[0.503, 0.636] | $0.792 \pm 0.014$<br>[0.760, 0.806] | $0.603 \pm 0.039$<br>[0.535, 0.656] | $0.607 \pm 0.043$<br>[0.532, 0.664] |
| A + B | $0.503 \pm 0.022$<br>[0.471, 0.549] | $0.786 \pm 0.013$<br>[0.760, 0.795] | $0.593 \pm 0.026$<br>[0.547, 0.638] | $0.596 \pm 0.027$<br>[0.548, 0.642] |
| A + C | $0.490 \pm 0.042$<br>[0.409, 0.582] | $0.735 \pm 0.022$<br>[0.718, 0.795] | $0.487 \pm 0.048$<br>[0.388, 0.552] | $0.477 \pm 0.057$<br>[0.358, 0.553] |
| A + B + C | $0.480 \pm 0.040$<br>[0.391, 0.543] | $0.770 \pm 0.017$<br>[0.743, 0.800] | $0.558 \pm 0.045$<br>[0.494, 0.636] | $0.557 \pm 0.052$<br>[0.482, 0.646] |

**Table S4:** Additional performance metrics of supervised metrics for the best-performing model type. The presented metrics concern the model type: tsfresh+XGBoost. All metrics are presented in the form: *mean ± std. deviation* [95% CI]

| Train Sites | Weighted $F_1$ | AUPRC |
| --- | --- | --- |
| B+C | $0.582 \pm 0.047$ [ 0.507, 0.688] | $0.714 \pm 0.024$ [0.675, 0.769] |
| A | $0.642 \pm 0.067$ [ 0.524, 0.737] | $0.809 \pm 0.042$ [0.750, 0.886] |
| A + B | $0.651 \pm 0.065$ [ 0.531, 0.747] | $0.830 \pm 0.055$ [ 0.726, 0.909] |
| A + C | $0.721 \pm 0.074$ [ 0.576, 0.822] | $0.845 \pm 0.038$ [ 0.767, 0.909] |
| A + B + C | $0.741 \pm 0.046$ [ 0.650, 0.814] | $0.886 \pm 0.035$ [0.819, 0.942] |

**Table S5:** Classification performance of unsupervised models. Training set composition varies by site. The test site was always site A, comprising both classes. Metrics reported: Balanced Accuracy and Macro *F*_1_. Values are reported as the mean and standard deviation along with the 95% confidence interval.

| Train Sites | Balanced Accuracy | Macro $F_1$ |
| --- | --- | --- |
| **tsfel** |  |  |
| B | $0.456 \pm 0.043$ [0.391, 0.525] | $0.449 \pm 0.045$ [ 0.375, 0.524] |
| C | $0.491 \pm 0.009$ [0.482, 0.500] | $0.414 \pm 0.004$ [ 0.409, 0.418] |
| **catch22** |  |  |
| B | $0.553 \pm 0.047$ [0.473, 0.650] | $0.549 \pm 0.049$ [ 0.472, 0.653] |
| C | $0.482 \pm 0.016$ [0.446, 0.500] | $0.409 \pm 0.008$ [ 0.391, 0.418] |
| **DTW+kNN** |  |  |
| B | $0.377 \pm 0.0$ [0.377, 0.377] | $0.382 \pm 0.0$ [ 0.382, 0.382] |
| C | $0.503 \pm 0.0$ [0.503, 0.503] | $0.500 \pm 0.0$ [ 0.500, 0.500] |
| **(Mini-)ROCKET** |  |  |
| B | $0.535 \pm 0.004$ [0.518, 0.536] | $0.293 \pm 0.009$ [ 0.259, 0.296] |
| C | $0.500 \pm 0.0$ [0.500, 0.500] | $0.418 \pm 0.0$ [ 0.418, 0.418] |

**Table S6:** Comparison of Model Performance to Human Baseline Using One-Sample Wilcoxon Tests. One-sample Wilcoxon tests were conducted to compare the performance of the tsfresh+XGBoost model against a human baseline (H_1_: model *>* human). The human baseline is defined as a logistic regression model fitted on the MDS-UPDRS III sum score. Δ denotes the mean difference (*x̄*_model_ *− x*_human_).

| Train Set | Metric | n | p-value | $\Delta$ |
| --- | --- | --- | --- | --- |
| A | AUROC | 30 | 0.9999 | -0.0667 |
| A | Precision at 80% Recall | 30 | 0.9972 | -0.0205 |
| A+B | AUROC | 30 | 0.9806 | -0.0437 |
| A+B | Precision at 80% Recall | 30 | 0.8252 | -0.0075 |
| A+C | AUROC | 30 | 0.6708 | -0.0091 |
| A+C | Precision at 80% Recall | 30 | 1.378e-03 | 0.0318 |
| A+B+C | AUROC | 30 | 8.708e-05 | 0.0578 |
| A+B+C | Precision at 80% Recall | 30 | 1.191e-04 | 0.0479 |

### S6 Defintion of Partial autocorrelation function

For a time series *T*={xt}_t=1_^n^, the partial autocorrelation at lag k, denoted as α*_k_*, quantifies the correlation between x*_t_* and x*_t−k_* after removing the linear influence of the intermediate observations x*_t−_*_1_,…,x*_t−k_*_+1_. It is defined as the partial correlation:

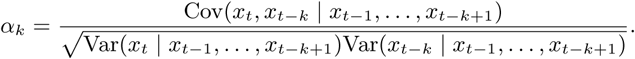

**Figure S3:**
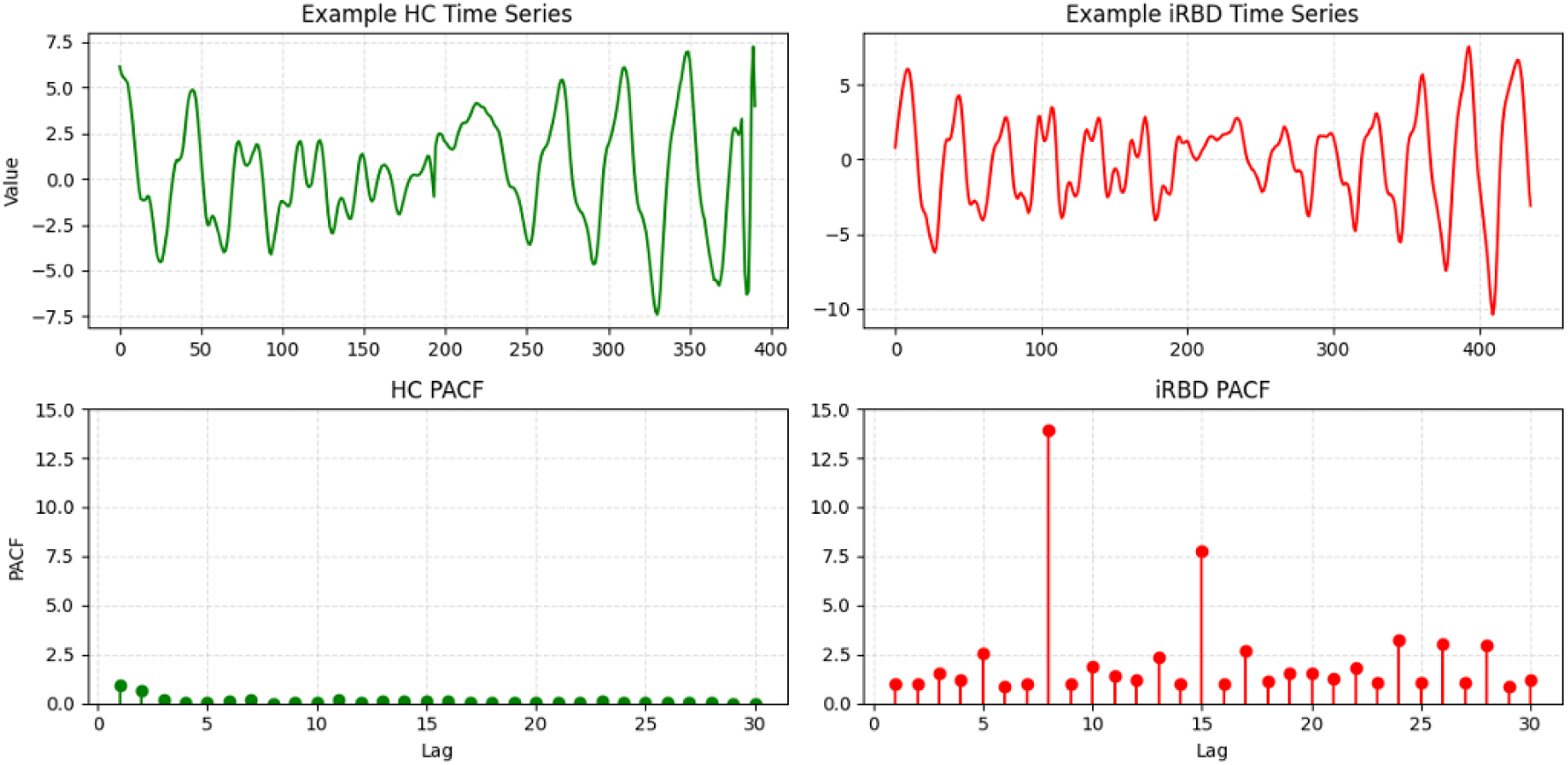
Illustration of Partial Autocorrelation Functions (PACFs). The top row shows two example time series extracted from the hip joints of participants in HC (left) and RBD (right) groups, with the x-axis indicating the frame index. The bottom row displays the corresponding absolute PACFs for lags 1-30.

**Table S7:** Bootstrap estimates of differences in model fit across striatal regions. Mean differences in the coefficient of determination (Δ*R*^2^) are reported together with their 95% bootstrap confidence intervals (CIs). Differences were calculated as PACF30 *−* MDS-UPDRS III, Combined *−* PACF30, and Combined *−* MDS-UPDRS III, respectively. Positive values therefore indicate greater explained variance for the first model in the corresponding comparison.

| Region | PACF30 vs. MDS-UPDRS III | Combined vs. PACF30 | Combined vs. MDS-UPDRS III |
| --- | --- | --- | --- |
| Caudate R | 0.067 [-0.166, 0.372] | 0.033 [0.000, 0.203] | 0.113 [0.001, 0.397] |
| Caudate L | -0.041 [-0.224, 0.203] | 0.057 [0.001, 0.259] | 0.056 [0.000, 0.271] |
| Anterior Putamen R | 0.119 [-0.139, 0.477] | 0.023 [0.000, 0.200] | 0.159 [0.000, 0.506] |
| Anterior Putamen L | 0.122 [-0.187, 0.494] | 0.019 [0.000, 0.216] | 0.150 [0.000, 0.513] |
| Posterior Putamen R | 0.151 [-0.161, 0.535] | 0.069 [0.000, 0.258] | 0.220 [0.001, 0.568] |
| Posterior Putamen L | -0.039 [-0.312, 0.398] | 0.222 [0.025, 0.388] | 0.183 [0.001, 0.536] |
| Putamen R | 0.092 [-0.204, 0.440] | 0.043 [0.000, 0.264] | 0.135 [0.000, 0.499] |
| Putamen L | -0.046 [-0.233, 0.299] | 0.101 [0.001, 0.292] | 0.055 [0.000, 0.400] |

**Table S8:** Top 30 tsfresh features ranked by SHAP importance. Features are ranked according to their mean absolute SHAP value across all test samples, i.e., participants from Site A. tsfresh descriptors capture complementary characteristics of gait dynamics, including temporal dependencies (e.g., autocorrelation and partial autocorrelation), frequency-domain properties (e.g., Fourier coefficients), localized time-frequency patterns (e.g., continuous wavelet transform coefficients), and signal complexity. The provided descriptions summarize the signal-level intuition of each feature and its corresponding movement characteristic.

| Feature | Intuition |
| --- | --- |
| diff_hips_partial_autocorrelation_lag_7 | Temporal dependency of hip movement after removing intermediate correlations. |
| dist_left_wrist-left_hip_cwt_coefficients__coeff_7__w_10__widths_(2,5,10,20) | Localized movement pattern of the left arm at a specific time scale. |
| diff_hips_partial_autocorrelation_lag_2 | Short-term temporal dependency of hip movement. |
| diff_shoulders_cwt_coefficients__coeff_6__w_10__widths_(2,5,10,20) | Localized oscillatory shoulder movement. |
| diff_shoulders_cwt_coefficients__coeff_3__w_10__widths_(2,5,10,20) | Transient shoulder movement at another frequency band. |
| angle_right_hip-(right_ankle-left_ankle)_fft_coefficient__attr_"imag"__coeff_62 | Phase-related frequency component of leg movement. |
| dist_right_wrist-right_hip_partial_autocorrelation_lag_7 | Long-range temporal dependency of right arm movement. |
| diff_shoulders_cwt_coefficients__coeff_1__w_2__widths_(2,5,10,20) | Very short-scale transient shoulder movement. |
| angle_left_hip-(left_ankle-right_ankle)_fft_coefficient__attr_"real"__coeff_25 | Magnitude of a specific movement frequency. |
| angle_left_hip-(left_ankle-right_ankle)_lempel_ziv_complexity_bins_3 | Complexity or irregularity of movement. |
| area_hips-ankles__energy_ratio_by_chunks__num_segments_10__segment_focus_6 | Fraction of movement energy occurring during a specific time interval. |
| diff_hips_partial_autocorrelation_lag_5 | Mid-range temporal dependency of hip movement. |
| dist_left_ankle-right_ankle__ratio_beyond_r_sigma_r_5 | Fraction of unusually large ankle separations. |
| diff_hips_cwt_coefficients__coeff_1__w_20__widths_(2,5,10,20) | Slow-changing component of hip movement. |
| angle_left_shoulder-(left_hip-left_wrist)_ratio_beyond_r_sigma_r_2.5 | Frequency of unusually large shoulder angles. |
| area_hips-ankles__energy_ratio_by_chunks__num_segments_10__segment_focus_5 | Movement energy concentrated in the middle of the recording. |
| angle_right_hip-(right_ankle-left_ankle)_partial_autocorrelation_lag_3 | Short-term dependency of leg orientation. |
| angle_right_hip-(right_ankle-left_ankle)_number_peaks_n_50 | Number of pronounced movement peaks. |
| dist_left_wrist-left_hip_fft_coefficient__attr_"imag"__coeff_39 | Phase-related frequency information of left arm movement. |
| angle_right_shoulder-(right_wrist-right_hip)_linear_trend__attr_"pvalue" | Significance of a linear movement trend over time. |
| dist_left_elbow-right_elbow_skewness | Asymmetry of elbow distance values. |
| area_hips-ankles__ratio_beyond_r_sigma_r_7 | Fraction of extreme body area values. |
| angle_left_shoulder-(left_hip-left_wrist)_autocorrelation_lag_3 | Similarity of shoulder movement after three time steps. |
| dist_left_knee-right_knee_fft_coefficient__attr_"abs"__coeff_34 | Magnitude of a specific knee movement frequency. |
| dist_left_knee-right_knee__ratio_beyond_r_sigma_r_1 | Fraction of knee distances outside one standard deviation. |
| angle_left_shoulder-(left_hip-left_wrist)_max_langevin_fixed_point_m_3_r_30 | Estimate of nonlinear movement stability. |
| diff_hips_fft_coefficient__attr_"imag"__coeff_83 | High-frequency phase component of hip movement. |
| dist_left_elbow-right_elbow_fft_coefficient__attr_"abs"__coeff_10 | Magnitude of a specific elbow movement frequency. |
| dist_left_elbow-right_elbow_partial_autocorrelation_lag_5 | Mid-range temporal dependency of elbow movement. |
| dist_left_elbow-right_elbow_max_langevin_fixed_point_m_3_r_30 | Estimate of nonlinear elbow movement stability. |

**Table S9:**
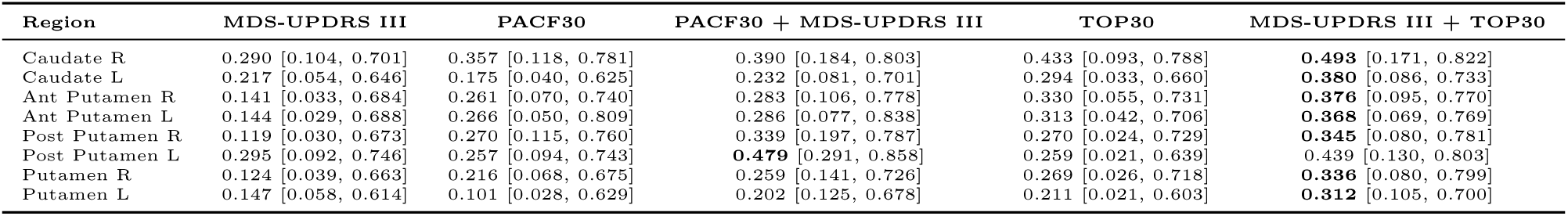
Model fit across striatal regions in terms of. *R*^2^. Values reflect the *R*^2^ from the model fitted on the full sample, with 95% bootstrap confidence intervals (CIs) shown in brackets. Models include MDS-UPDRS III only, PACF30 only, full combined (PACF30 + MDS-UPDRS III), top-30 gait features, and MDS-UPDRS III + top-30 gait features. Higher values indicate better predictive performance.

**Table S10:** Model fit across striatal regions in terms of AIC. Values reflect the AIC from the model fitted on the full sample, with 95% bootstrap confidence intervals (CIs) shown in brackets. Models include MDS-UPDRS III only, PACF30 only, full combined (PACF30 + MDS-UPDRS III), top-30 gait features, and MDS-UPDRS III + top-30 gait features. Lower values indicate better model fit.

| Region | MDS-UPDRS III | PACF30 | PACF30 + MDS-UPDRS III | TOP30 | MDS-UPDRS III + TOP30 |
| --- | --- | --- | --- | --- | --- |
| Caudate R | 15.441 [-10.827, 23.110] | 13.256 [-14.206, 23.529] | 14.080 [-13.728, 23.158] | 7.612 [-16.612, 23.837] | **7.065** [-17.483, 22.458] |
| Caudate L | 9.138 [-13.459, 17.663] | 10.272 [-12.435, 18.724] | 10.699 [-15.056, 18.232] | 4.747 [-13.914, 18.752] | **3.633** [-16.410, 17.762] |
| Ant Putamen R | 19.272 [-7.104, 26.908] | 16.581 [-10.692, 23.711] | 18.022 [-10.266, 23.950] | **12.169** [-11.659, 25.734] | 12.737 [-11.586, 26.264] |
| Ant Putamen L | 14.743 [-6.786, 20.031] | 11.974 [-16.601, 19.036] | 13.491 [-17.316, 19.851] | **8.690** [-7.578, 19.925] | 8.950 [-10.538, 21.216] |
| Post Putamen R | 23.173 [-9.729, 32.776] | 19.591 [-12.284, 27.442] | 19.703 [-13.400, 25.590] | 16.133 [-11.588, 34.136] | **15.863** [-17.077, 32.944] |
| Post Putamen L | 18.774 [-9.896, 28.941] | 19.791 [-8.590, 26.344] | 15.052 [-18.285, 20.569] | 16.327 [-6.373, 32.320] | **12.436** [-15.605, 29.697] |
| Putamen R | 19.282 [-12.053, 29.080] | 17.067 [-13.050, 25.034] | 17.941 [-13.332, 24.667] | 12.710 [-17.295, 29.613] | **12.564** [-21.426, 29.507] |
| Putamen L | 14.167 [-12.622, 23.950] | 15.274 [-13.071, 21.529] | 14.778 [-14.050, 19.763] | 9.912 [-14.711, 24.677] | **8.904** [-17.787, 23.971] |

**Table S11:** Model fit across striatal regions in terms of corrected AIC (AICc). Values reflect the AICc from the model fitted on the full sample, with 95% bootstrap confidence intervals (CIs) shown in brackets. Models include MDS-UPDRS III only, PACF30 only, full combined (PACF30 + MDS-UPDRS III), top-30 gait features, and MDS-UPDRS III + top-30 gait features. Lower values indicate better model fit.

| Region | MDS-UPDRS III | PACF30 | PACF30 + MDS-UPDRS III | TOP30 | MDS-UPDRS III + TOP30 |
| --- | --- | --- | --- | --- | --- |
| Caudate R | 19.191 [-10.827, 23.110] | 17.006 [-14.206, 23.529] | 19.680 [-13.728, 23.158] | **11.362** [-12.862, 27.587] | 12.665 [-11.883, 28.058] |
| Caudate L | 12.888 [-13.459, 17.663] | 14.022 [-12.435, 18.724] | 16.299 [-15.056, 18.232] | **8.497** [-10.164, 22.502] | 9.233 [-10.810, 23.362] |
| Ant Putamen R | 24.272 [-7.104, 26.908] | 21.581 [-10.692, 23.711] | 25.659 [-10.266, 23.950] | **17.169** [-6.659, 30.734] | 20.374 [-3.949, 33.900] |
| Ant Putamen L | 19.743 [-6.786, 20.031] | 16.974 [-16.601, 19.036] | 21.127 [-17.316, 19.851] | **13.690** [-2.578, 24.925] | 16.586 [-2.901, 28.853] |
| Post Putamen R | 27.788 [-9.729, 32.776] | 24.207 [-12.284, 27.442] | 26.703 [-13.400, 25.590] | **20.748** [-6.972, 38.751] | 22.863 [-10.077, 39.944] |
| Post Putamen L | 23.390 [-9.896, 28.941] | 24.406 [-8.590, 26.344] | 22.052 [-18.285, 20.569] | 20.943 [-1.758, 36.935] | **19.436** [-8.605, 36.697] |
| Putamen R | 23.568 [-12.053, 29.080] | 21.353 [-13.050, 25.034] | 24.402 [-13.332, 24.667] | **16.995** [-13.010, 33.898] | 19.025 [-14.964, 35.968] |
| Putamen L | 18.167 [-12.622, 23.950] | 19.274 [-13.071, 21.529] | 20.778 [-14.050, 19.763] | **13.912** [-10.711, 28.677] | 14.904 [-11.787, 29.971] |

